# Costs and cost-effectiveness of interventions based on third generation therapy models to treat symptoms of depression and anxiety: A systematic review and meta-analysis

**DOI:** 10.1101/2025.05.05.25327011

**Authors:** Yscenia Paredes-Gonzales, Gianfranco Centeno-Terrazas, Kelly De la Cruz-Torralva, Elisa Romani-Huacani, Yan Pieer Alexis-Montalban Lozada, David Villarreal-Zegarra

## Abstract

**Background:** This study aimed to evaluate the costs and cost-effectiveness of third-generation therapies for treating depressive and anxiety symptoms.

**Methods:** We conducted a systematic review and meta-analysis, searching SCOPUS, Web of Science, PsycINFO, EMBASE, and MEDLINE. Randomized controlled trials reporting costs or cost-effectiveness of third-generation therapies for depression or anxiety were included. Meta-analysis of quality-adjusted life years (QALYs) was conducted using Hedges’ g and random-effects models. Costs were standardized to US dollars and adjusted for inflation.

**Results:** Twenty studies met inclusion criteria, with 10 suitable for meta-analysis. Meta-analysis showed no significant difference in QALY improvement between third-generation therapies and control conditions (Hedges’ g=0.015, 95%CI: -0.001 to 0.032; P=0.070; I²=47.8%). Subgroup analyses by therapy type (Mindfulness-Based Cognitive Therapy, Behavioral Activation, Acceptance and Commitment Therapy, and Problem-Solving Therapy) also yielded non-significant results. High risk of bias was observed in 50% of studies due to narrow perspective, in 45% due to omission of cost measurement, and in 46% due to insufficient information on discounting. Publication bias was detected (Egger’s test: P=0.011). Our study found no significant difference in the average cost per person between receiving an intervention based on third-generation therapies (M=$2,494.6; SD=$7,090.9) and receiving a control condition (M=$1,912.0; SD=$2,081.3) (t=0.253; p=0.802).

**Conclusions:** Our study found that interventions based on third-generation therapies do not entail significant additional costs compared to control conditions. However, the interventions based on third-generation therapies did not improve the QALYs of the participants more than the control group.

## Background

As reported by the World Health Organization (WHO), the global estimated prevalence of depression is 4.4%, with anxiety representing an additional 3.6% [1]. These disorders not only diminish individuals’ productivity and economic capabilities but also cause profound human and social suffering [2,3]. Depression is the leading cause of disability worldwide, affecting approximately 322 million individuals and accounting for 7.5% of total disability-adjusted life years [1]. Furthermore, major depression and anxiety disorder are among the primary contributors to the global burden of disease [4,5].

It is estimated that the global cost of mental disorders, including depression and anxiety, will reach $6 trillion by 2030 [6]. Furthermore, depression and anxiety are markedly linked to diminished workplace productivity, resulting in elevated rates of absenteeism and presenteeism [7,8]. These indirect costs, in conjunction with the direct costs of treatment, underscore the necessity for the implementation of efficacious and economically sustainable interventions.

In response to these mental health challenges, third-generation therapies have emerged as a promising alternative. These include Acceptance and Commitment Therapy (ACT), Dialectical Behavior Therapy (DBT), Compassion Therapy, Behavioral Activation, and Mindfulness-Based Therapy, among others. These approaches were developed over the past few decades and emphasize the functionality of thoughts and emotions rather than their frequency or form [9–11].

The efficacy of these therapies in the treatment of depression and anxiety has been repeatedly demonstrated in scientific research. A meta-analysis revealed that ACT is an efficacious treatment for these conditions, demonstrating improvements in both symptom severity and overall quality of life [12]. Another study demonstrated that DBT is particularly efficacious for severe anxiety and depressive disorders, significantly enhancing emotional regulation [13]. Behavioral Activation has been demonstrated to be as efficacious as antidepressant medications for the treatment of severe depression [14]. Additionally, Mindfulness-Based Therapy emphasizes the importance of mindfulness, defined as the state of deliberate and conscious attention to the present moment, observing and accepting experiences as they are without issuing critical or judgmental evaluations [15]. These findings underscore the potential of these interventions to effectively address symptoms of anxiety and depression, offering therapeutic alternatives supported by a robust scientific foundation.

Nevertheless, despite the growing body of evidence supporting the efficacy of these therapies, information on their cost-effectiveness remains limited. Economic evaluations of mental health interventions are of critical importance for the formulation of health policy and the efficient allocation of resources [16,17]. Cost-effectiveness analyses are employed to ascertain whether an intervention offers optimal value for money, comparing its costs with the benefits it delivers, typically quantified in quality-adjusted life years (QALYs) [18].

In health systems with constrained resources, it is of paramount importance to identify interventions that offer the greatest benefits at the lowest costs [19]. Furthermore, cost-effectiveness considerations can influence the adoption and dissemination of new therapies, directly impacting patient access to potentially beneficial treatments [20].

To date, there have been few systematic reviews that have focused on the cost-effectiveness of third-generation therapies for depression and anxiety, and those that have been conducted have had limited scope. A review conducted in 2018 identified 11 studies that evaluated the economic impact of these therapies [21]. However, the aforementioned review did not provide a comprehensive cost analysis or a specific assessment of the risk of bias in the economic evaluations. The rapid evolution of the field of mental health and the increasing use of these therapies underscore the need for an updated review of the available evidence.

The absence of an up-to-date and comprehensive synthesis of the evidence on the cost-effectiveness of third-generation therapies represents a notable gap in the existing literature. Such information is vital for public health decision-makers, healthcare providers, and researchers seeking to optimize resources and enhance outcomes in the treatment of depression and anxiety.

In this context, this systematic review aims to evaluate the current evidence on the costs and cost-effectiveness of interventions based on third-generation models for the treatment of depression and anxiety symptoms.

## Methods

### Protocol and Registration

This study, registered with PROSPERO (CRD42024539215), was conducted in accordance with the PRISMA guidelines for systematic reviews, thereby ensuring that the data collection and presentation were transparent, comprehensive, and accurate (see supplementary material 1).

Our study presents a deviation from protocol, our study initially aimed to evaluate cost-effectiveness from a comprehensive perspective, considering improvements in quality of life, QALYs, and reduction of symptoms of depression and anxiety. However, due to the complexity of the search process, we narrowed our focus to a cost-effectiveness evaluation based solely on QALYs.

### Inclusion Criteria

The eligibility criteria were defined in accordance with the PICO framework, aiming to include the general population without any age restrictions, encompassing both individuals without comorbidities and those with physical or mental health conditions. The study focused on third-generation therapies and interventions delivered through various modalities, including in-person consultations, guided teleconsultations, synchronous care through videoconferencing, and self-guided interventions. These interventions were provided by healthcare professionals, technicians, and non-specialists or implemented as self-guided interventions without support from healthcare professionals. Active comparators considered included psychotherapy, psychoeducation, pharmacotherapy, usual care, bibliotherapy, animal-assisted therapy, among others. Passive comparators considered included waiting lists, no treatment, or placebo. The primary outcomes of interest were the costs and cost-effectiveness of these interventions for the treatment of depressive and anxiety symptoms. Costs were defined as the financial resources required to implement an intervention, including direct costs for materials, personnel, technology, and infrastructure [18]. Cost-effectiveness was understood as the ratio between the costs of an intervention and its benefits in terms of quality-adjusted life years (QALYs), a unit of measurement that combines both the quantity and quality of life, used to assess a person’s quality of life after a health intervention [22].

The study included only randomized controlled trials. Documents that constituted reviews, protocols, theses, or any other materials that had not undergone peer review were excluded from the review. The inclusion period ranged from the advent of third-generation therapies in 1980 to March 12, 2024 (the search date and first extracted). No language restrictions were applied.

### Sources of Information and Search Strategy

A variety of databases were utilized, including SCOPUS, Web of Science, PsycINFO, EMBASE, and MEDLINE. Each database was searched using a tailored strategy incorporating specific keywords related to third-generation therapies, costs, cost-effectiveness, clinical trials, depression, and anxiety (see supplementary material 2).

### Selection of Evidence Sources

The selection process was conducted in three stages. Initially, studies identified from the search were downloaded in RIS format. They were then imported into an EndNote X9 file, where duplicates were removed using both automatic and manual methods available in EndNote X9.

Next, duplicate-free records were exported to a RIS file to begin the title and abstract screening process. This was performed using ASReview, a free software that leverages artificial intelligence to streamline the review by prioritizing documents most likely to meet our inclusion criteria. ASReview’s Oracle mode, Naive Bayes classifier, and TF-IDF feature extraction technique were employed (https://asreview.nl/). Title and abstract screening was carried out by two independent reviewers, with conflicts resolved through discussion. In cases of persistent disagreement, a third reviewer made the final decision on inclusion or exclusion of records.

Finally, full-text reviews were conducted using the Rayyan AI platform (https://www.rayyan.ai/), where two independent reviewers assessed all documents that passed the title and abstract review. In case of disagreements, a third reviewer had the final say on the inclusion or exclusion of studies. Excluded documents were listed, and reasons for exclusion were detailed in supplementary material.

### Data Extraction

Two reviewers groups independently extracted relevant data using a custom Excel extraction form (GCT and KDCT; ERH and YPAML). Any discrepancies were discussed between the reviewers to reach a consensus. If conflicts persisted, a third reviewer made the final decision on the data in question (YPG). When the necessary data for meta-analysis were not available, the authors of the respective studies were contacted to request the required information.

Information collected included publication details (title, first author’s surname, publication year, and country of the first author’s affiliation), study design characteristics (number of groups in the clinical trial, study type, etc.), participant details (percentage of women, total number of participants, population type, and a brief sample description), and intervention characteristics (type of third-generation therapy used, delivery mode, frequency of use, number of sessions, and a detailed description). Control group characteristics were specified, including type (active or passive) and a description. Depressive symptoms measurement was detailed through the instrument used and measurement method. Data on costs and cost-effectiveness and other relevant aspects were also collected. Finally, specific study characteristics such as conflicts of interest and funding sources were documented.

### Critical Appraisal of Risk of Bias in the Evidence Sources

The ECOBIAS checklist [23] was used to assess the risk of bias in economic evaluations. This tool was chosen for its specificity in health economic evaluations and its ability to identify potential biases in such studies. It focused on eleven areas: 1) Narrow perspective bias, 2) Inefficient comparator bias, 3) Omission of cost measurement bias, 4) Intermittent data collection bias, 5) Invalid valuation bias, 6) Ordinal ICER bias, 7) Double counting bias, 8) Inappropriate discounting bias, 9) Limited sensitivity analysis bias, 10) Sponsor bias, and 11) Reporting and dissemination bias. A study was considered methodologically robust if it scored well on the ECOBIAS checklist.

### Analysis plan

#### Characteristics of the studies

A narrative description was provided for each study included in our review, detailing their individual characteristics and findings. Additionally, we presented the cost and cost-effectiveness information from each study.

#### Cost analysis

To compare economic outcomes across studies, costs were standardized to U.S. dollars (USD) using the average exchange rate for the year of each study, as reported by the World Bank (https://data.worldbank.org/indicator/PA.NUS.FCRF). Costs were further adjusted for the latest annual inflation rate reported by the World Bank (https://datos.bancomundial.org/indicator/FP.CPI.TOTL.ZG) to ensure comparability across studies. The exchange rates and inflation rates used are detailed in the corresponding tables.

We calculated the average cost per person across all studies. This was based on the reported average cost-per-person in each study, multiplied by the total number of participants. Total costs were summed separately for intervention and control groups. These total costs were then divided by the total number of participants in each respective group, yielding the average cost-per-person across all studies included in the review.

Following Planas-Miret et al. [24], we categorized costs into nine main types: direct medical costs, direct non-medical costs, indirect costs (covering absenteeism and presenteeism), opportunity costs, informal care costs (including those provided by family and friends), social costs, rehabilitation costs, and home care costs.

For this study, we reported based on the types of costs reported, structuring them into four categories: first, direct medical costs; second, direct non-medical costs; third, indirect costs; and lastly, social costs.

It is important to note that the CHEERS 2022 statement is intended to be used for different forms of health economic evaluation. These include analyses that only examine costs and cost trade-offs (i.e. cost analysis) or those that examine both costs and consequences. The latter include analyses that take into account health consequences (such as cost-effectiveness/utility analyses (CEA/CUA), cost-minimization, cost-benefit/cost-benefit analyses (CBA)) and broader measures of benefit and harm to individuals (such as extended CEA/CBA), including equity measures (such as distributional CEA). While we recognize that some studies that compare costs are labeled as CBA, we recommend using this term for studies that include a monetary valuation of health outcomes

An independent mean difference analysis was performed to compare the average cost per person between the intervention and control groups. This analysis was based on all studies that provided average cost-per-person data for either the control or intervention group. In addition, a subanalysis was performed focusing only on those studies that reported costs for both groups.

#### Meta-analysis of Cost-Effectiveness Analysis

For the cost-effectiveness analysis, we conducted a meta-analysis of the studies that reported sufficient data on Quality-Adjusted Life Years (QALYs) using STATA version 18 software. We used Hedges’ g as the effect size measure to account for potential small sample bias. The difference in QALYs between the intervention and control groups was calculated by subtracting the control group QALYs from the intervention group QALYs. A random-effects model was employed for the meta-analysis due to the expected heterogeneity among studies. We assessed heterogeneity using the I² statistic, with values of 25%, 50%, and 75% indicating low, moderate, and high heterogeneity, respectively. The overall effect size and its 95% confidence interval were calculated, and statistical significance was set at p < 0.05. We also conducted subgroup analyses based on the type of third-generation therapy. The therapies analyzed included Mindfulness-Based Cognitive Therapy (MBCT), Behavioral Activation and Mindfulness, Behavioral Activation, Acceptance and Commitment Therapy (ACT), and Problem-Solving Therapy. For each subgroup, we calculated the effect size, 95% confidence interval, and assessed heterogeneity.

Forest plots were generated to visually represent the results of both the overall meta-analysis and the subgroup analyses. These plots display the effect sizes and confidence intervals for individual studies and the pooled estimates, allowing for easy interpretation of the results and assessment of between-study variability. The assessment of publication bias was performed using Egger’s regression test and visual inspection of the distribution of studies in the funnel plot. This was carried out since the meta-analysis included at least 10 studies, which ensures adequate capacity to detect real asymmetries [25]. Publication bias was considered to be present if the distribution of studies in the funnel plot was asymmetric and if the result of Egger’s test was significant (p < 0.05).

## Result

### Study Selection

Initially, a total of 2,981 records were identified through five databases and registries. Of these, 1,260 were eliminated as duplicates, reducing the number of records to 1,721. Subsequently, a review by title and abstract was conducted, excluding 1,624 records for not meeting the inclusion criteria, which left 97 records for full-text review. The full-text review resulted in the exclusion of 77 additional records. The reasons for exclusion for each record during the full-text evaluation can be found in supplementary material 3. Finally, 20 studies met all inclusion criteria and were selected for inclusion in the systematic review (see Figure 1).

**Figure 1.**
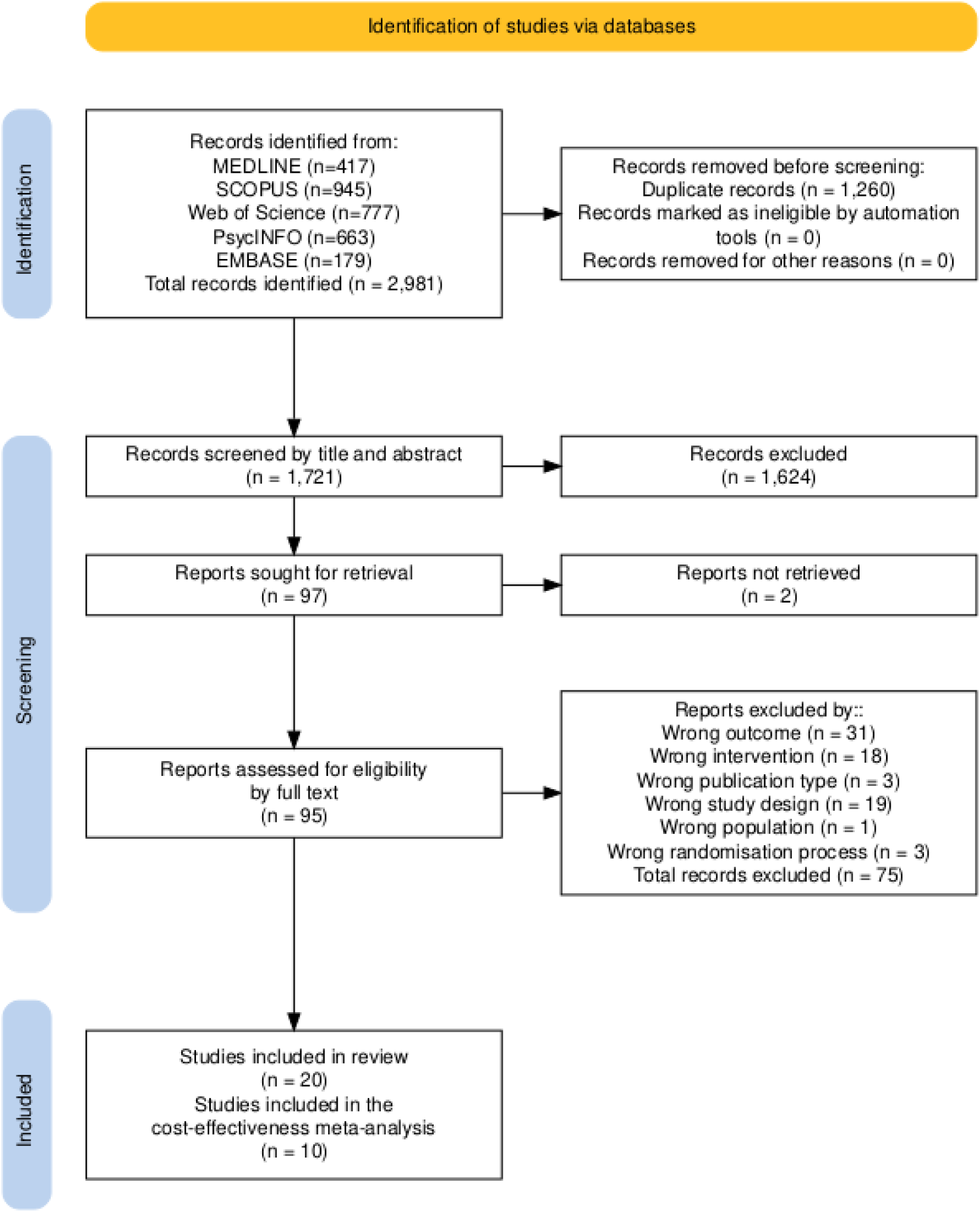
Flowchart.

### Study Characteristics

The systematic review included 20 studies [26–45]. Most studies were conducted between 2010 and 2019 (n=10; 50%). Almost all studies were published in English (n=19; 95%), and the country where most studies were conducted was the United Kingdom (n=8; 40%). Also, 95% of the studies were conducted in high-income countries (n=19). At the clinical trial level, 70% of them used a superiority framework (n=14), all studies had a parallel design, 80% of the studies had only two arms (intervention and control; n=16), and 80% of the studies had a duration of between 8 to 12 weeks (n=16). In addition, 65% of the studies were conducted in populations with mental health problems. We did not find any studies evaluating the cost-effectiveness or costs of teleconsultation, mHealth interventions or self-guided intervention that meet the inclusion criteria.

Regarding the evaluated outcomes, 85% assessed depressive symptoms (n=17), 50% assessed depressive symptoms (n=10), and 85% evaluated quality-adjusted life years or QALYs (n=17). Additionally, 90% of the studies conducted a cost analysis (n=18) and only 65% of the included studies reported cost-effectiveness (n=13). The most commonly used third-generation therapies as interventions were mindfulness-based cognitive therapy (n=7; 35%), acceptance and commitment therapy (n=6; 30%), and behavioral activation (n=5; 25%). On the other hand, the most used control was treatment as usual (n=9; 45%).

The characteristics of the included studies can be seen in Table 1. The individual description of each of the studies included in the systematic review is presented in supplementary material 4.

**Table 1.** Characteristics of the Included Studies (n=20).

|  | n | % |
| --- | --- | --- |
| **Publication year** |  |  |
| 2000-2009 | 1 | 5% |
| 2010-2019 | 10 | 50% |
| 2020-2024 | 9 | 45% |
| **Article language** |  |  |
| German | 1 | 5% |
| English | 19 | 95% |
| **Study country** |  |  |
| Germany | 1 | 5% |
| Australia | 1 | 5% |
| China | 1 | 5% |
| Denmark | 1 | 5% |
| Spain | 1 | 5% |
| United States | 1 | 5% |
| Netherlands | 2 | 10% |
| United Kingdom | 8 | 40% |
| Sweden | 4 | 20% |
| **Country income level** |  |  |
| Upper middle-income economies | 1 | 5% |
| High-income economies | 19 | 95% |
| **Study framework** |  |  |
| Superiority | 14 | 70% |
| Non-inferiority | 2 | 10% |
| Unclear | 4 | 20% |
| **Study design** |  |  |
| Parallel | 20 | 100% |
| **Number of study arms** |  |  |
| 2 | 16 | 80% |
| 3 | 2 | 10% |
| 4 | 2 | 10% |
| **Type of intervention\*** |  |  |
| Acceptance and Commitment Therapy (ACT) | 6 | 23.08% |
| Dialectical Behavioral Therapy (DBT) | 1 | 3.85% |
| Behavioral Activation | 5 | 19.23% |
| Mindfulness-Based Cognitive Therapy (MBCT) | 8 | 30.77% |
| Behavioral activation and mindfulness | 1 | 3.85% |
| Workplace dialog intervention (WDI) and Acceptance and Commitment Therapy (ACT) | 2 | 7.69% |
| Workplace dialog intervention (WDI) | 2 | 7.69% |
| Problem solving therapy | 1 | 3.85% |
| **Type of Control\*** |  |  |
| Waiting List | 5 | 19.23% |
| Usual Treatment | 12 | 46.15% |
| Cognitive behavioral therapy | 3 | 11.54% |
| Self-help | 3 | 11.54% |
| Applied relaxation | 1 | 3.85% |
| Telephone support calls | 2 | 7.69% |
| **Delivery modality\*** |  |  |
| Guided by a health professional | 22 | 84.62% |
| Guided with self-applied components | 4 | 15.38% |
| **Duration in weeks** |  |  |
| 4 | 2 | 10% |
| 6 | 1 | 5% |
| 8 | 9 | 45% |
| 12 | 7 | 35% |
| 16 | 1 | 5% |
| **Types of population** |  |  |
| Elderly population | 2 | 10% |
| Population with mental health problems | 13 | 65% |
| Women-only population | 1 | 5% |
| Population with chronic diseases | 4 | 20% |
| **Reports depressive symptoms** |  |  |
| Not evaluated | 3 | 15% |
| Main outcome | 14 | 70% |
| Secondary outcome | 3 | 15% |
| **Reports anxious symptoms** |  |  |
| Not evaluated | 10 | 50% |
| Main outcome | 5 | 25% |
| Secondary outcome | 5 | 25% |
| **Reports QALYs** |  |  |
| No | 3 | 15% |
| Yes | 17 | 85% |
| **Reports costs** |  |  |
| No | 2 | 10% |
| Yes | 18 | 90% |
| **Reports other types of cost-effectiveness** |  |  |
| No | 7 | 35% |
| Yes | 13 | 65% |
| **Currency type** |  |  |
| U.S. dollar | 5 | 25% |
| Euro | 7 | 35% |
| Pound Sterling | 7 | 35% |
| Australian dollar | 1 | 5% |
| **Conflict of interest** |  |  |
| No | 11 | 55% |
| Potential | 1 | 5% |
| Not reported | 8 | 40% |
| **Funding** |  |  |
| Not reported | 7 | 35% |
| Corporations | 2 | 10% |
| Government | 10 | 50% |
| Self-financed | 1 | 5% |

Note: n = number, % = percentage. QALYs = quality-adjusted life years. \* Studies with multiple arms are considered (n = 26).

### Risk of Bias in Individual Studies

Figure 2A shows the risk of bias for each individual study. The most robust studies found were those by Richards et al. (2016) [38] and Sanjib Saha et al. (2020) [40]. Figure 2B presents the proportion of responses within ECOBIAS (Low risk of bias, some concerns, high risk of bias, and not reported) only for those studies where each of the eleven ECOBIAS criteria was applicable. It was observed that 50% of the studies presented a high risk of bias due to narrow perspective, indicating that these studies evaluated costs from a limited perspective, potentially omitting significant costs and outcomes. Additionally, 45% of the studies reported a high risk of bias due to omission of cost measurement (n=9), which implied the omission of relevant costs, resulting in an overestimation or underestimation of an intervention’s profitability. Furthermore, 45% of the studies did not report sufficient information to assess bias due to inappropriate discounting (n=9), as they did not mention discount rates or provided limited or incorrect information about them, which could significantly alter the perception of an intervention’s profitability.

**Figure 2.**
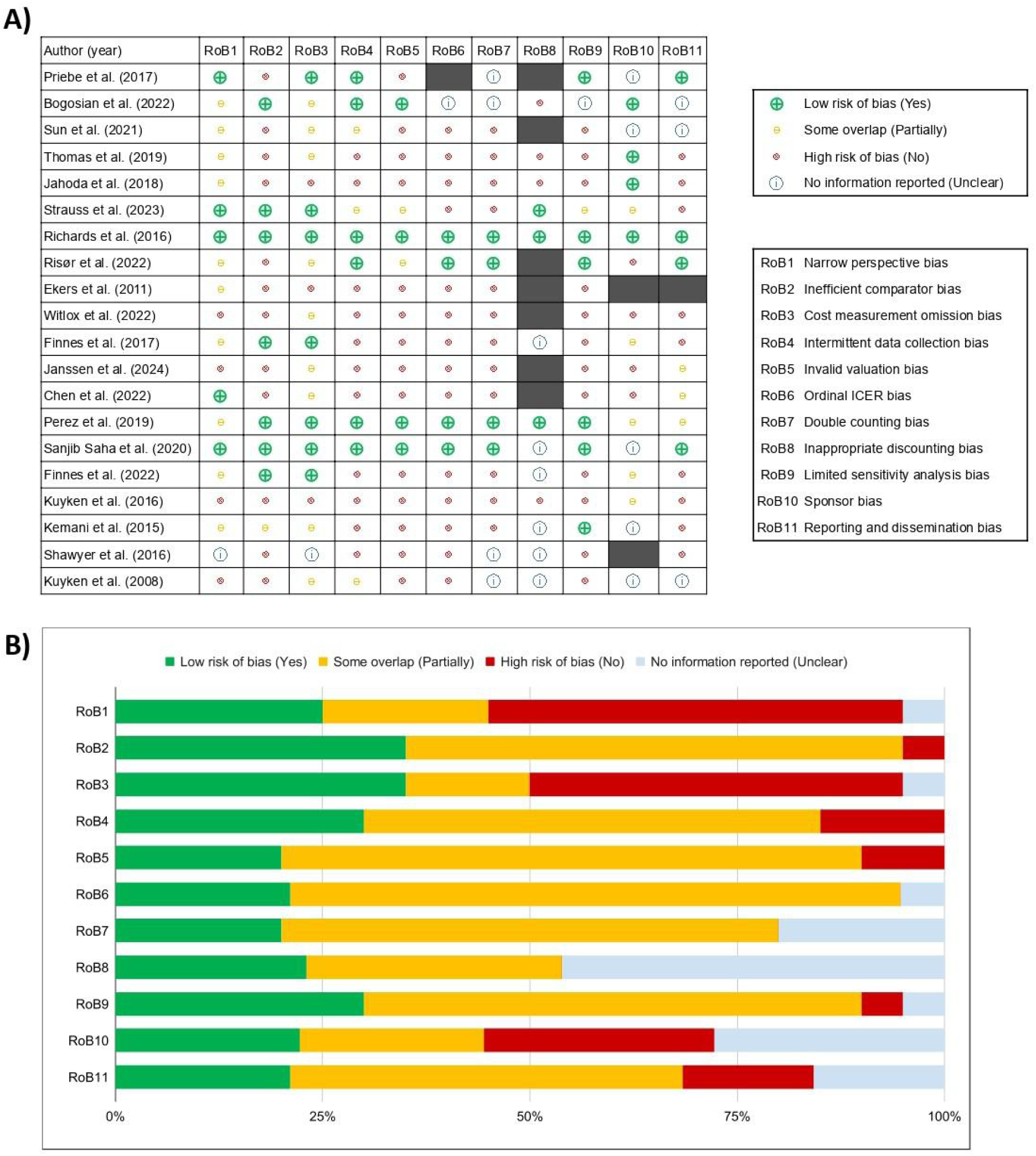
Risk of bias assessment using ECOBIAS (n = 20). Note: A) Individual risk of bias. In dark gray are the cases where the RoB criterion is applicable. B) Pooled risk of bias, for those that apply.

### Cost analysis

In the cost analysis, four different types of costs were identified (direct medical costs, direct non-medical costs, indirect costs, and social costs) reported within the included studies (see Table 2). The types of costs most frequently reported in the included studies were mental health care costs (n=17; 85%) and mental health medical specialist costs (n=15; 75%). Potentially, these costs were the most reported as the studies focused on mental health. On the other hand, 60% of the studies (n=12) reported costs for hospital care, medication costs, and absenteeism costs. The types of costs that were reported less frequently were rehabilitation costs (n=2; 10%), costs of using therapeutic devices (n=2; 10%), informal care costs (n=2; 10%), and domestic help costs (n=1; 5%). It should be noted that no study reported the opportunity cost referring to costs generated for users by ceasing productive activities, such as being unable to work due to waiting to receive medical care.

**Table 2.**
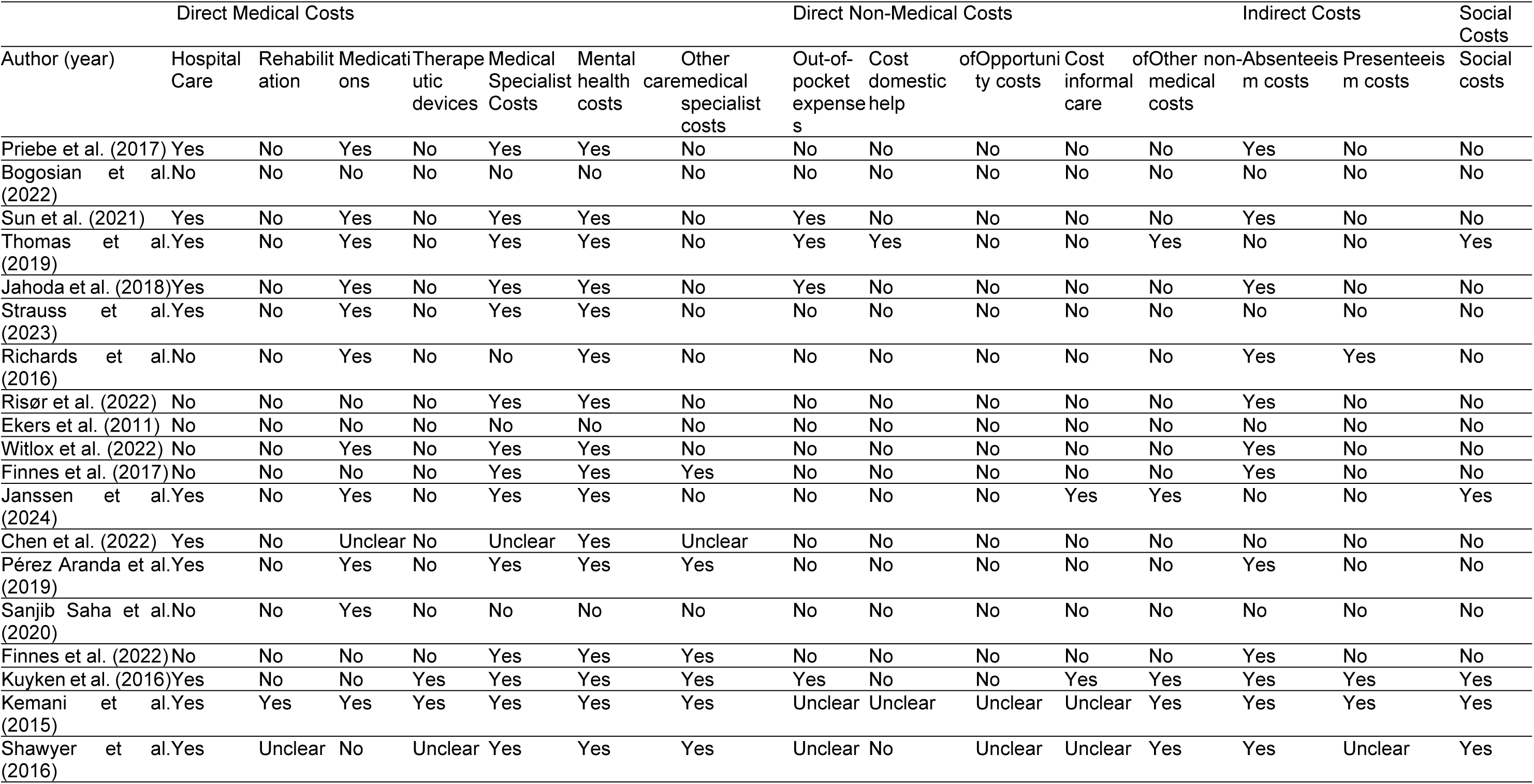
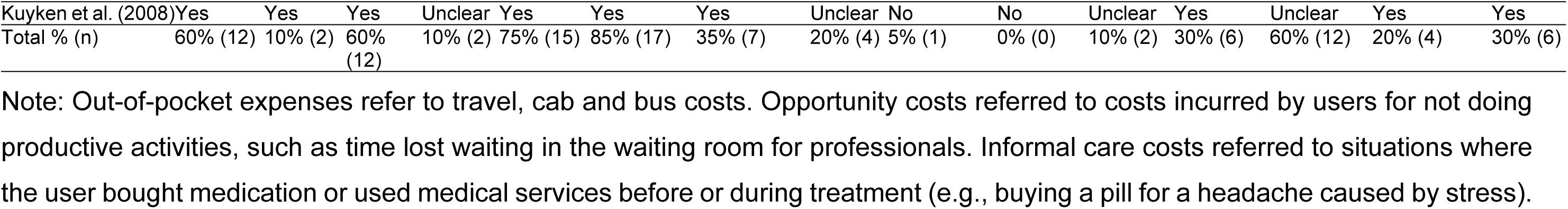
Types of costs reported in the included studies (n=20).

Our study identified direct medical, non-medical, indirect, social, and per-person costs for each of the studies in the currency types reported in them (see Table 3). However, these amounts were not comparable because they did not consider the exchange rate or annual inflation. Therefore, an average cost per person analysis was performed based on those studies that reported this information.

**Table 3.**
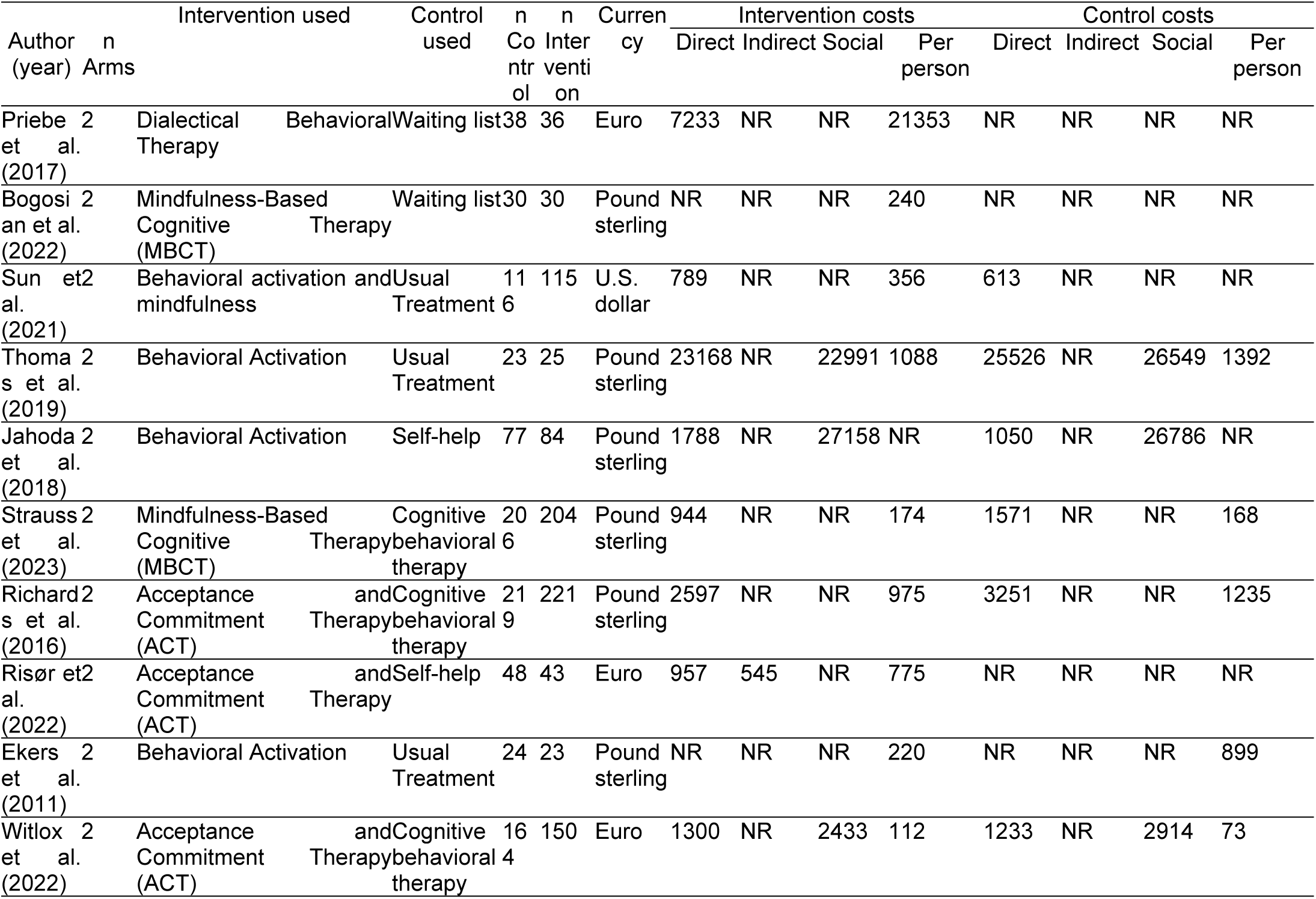
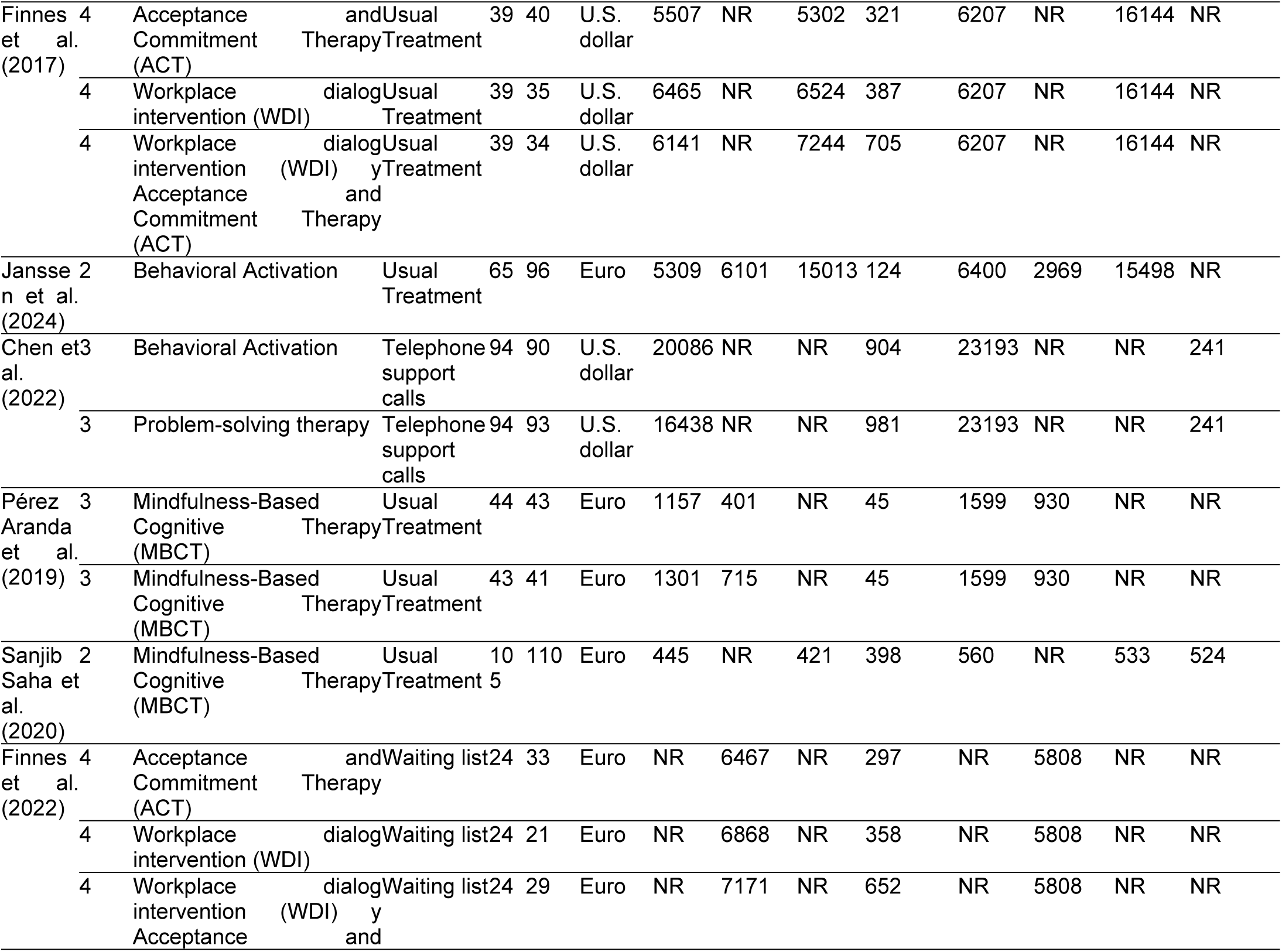
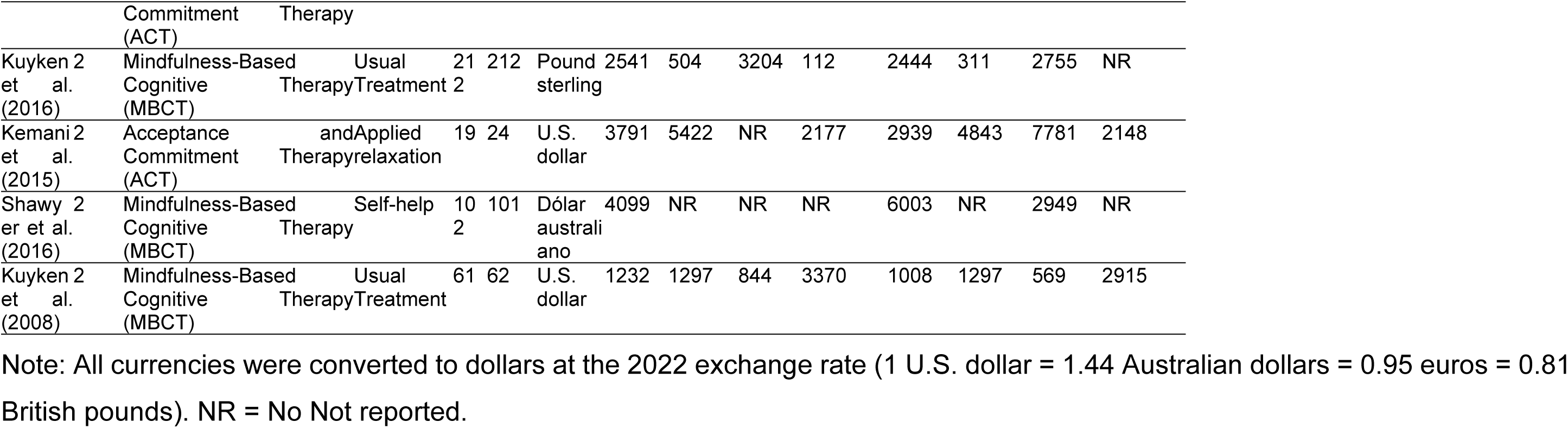
Direct, indirect, social and per person costs for each of the intervention and control arms (n=26).

Table 4 presents the results of the cost per person analyses for the different interventions and controls of the different arms (n=26), including studies with three and four arms. In 92.3% of cases, cost per person information was reported for the intervention groups (n=24) and only in 38.5% of cases were costs per person reported for the control groups (n=10). Our study found that there was no significant difference in the average cost per person between the groups receiving an intervention based on third-generation therapies (M = $2,494.6; SD = $7,090.9) and those assigned to a control condition (M = $1,912.0; SD = $2,081.3) (t = 0.253; p=0.802). In addition, a subanalysis was performed that included only those studies that reported average costs per person in both the intervention (M = $1,896.0; SD = $2,293.2) and control groups (M = $1,912.0; SD = $2,081.3) and again found no significant differences between the two groups (t = -0.016; p=0.987). These results suggest that an intervention based on third-generation therapies does not result in significant additional costs compared to control conditions.

**Table 4.**
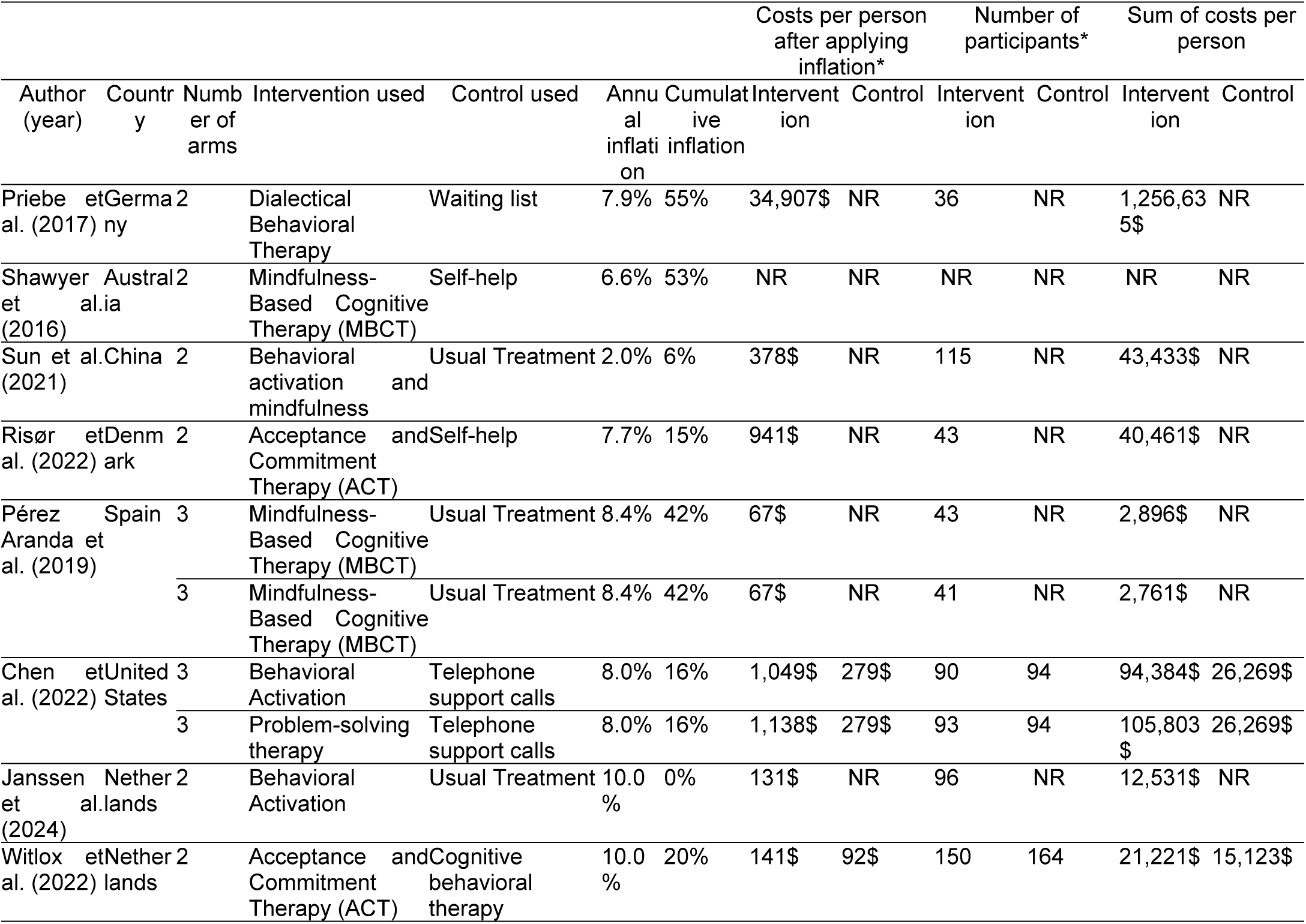
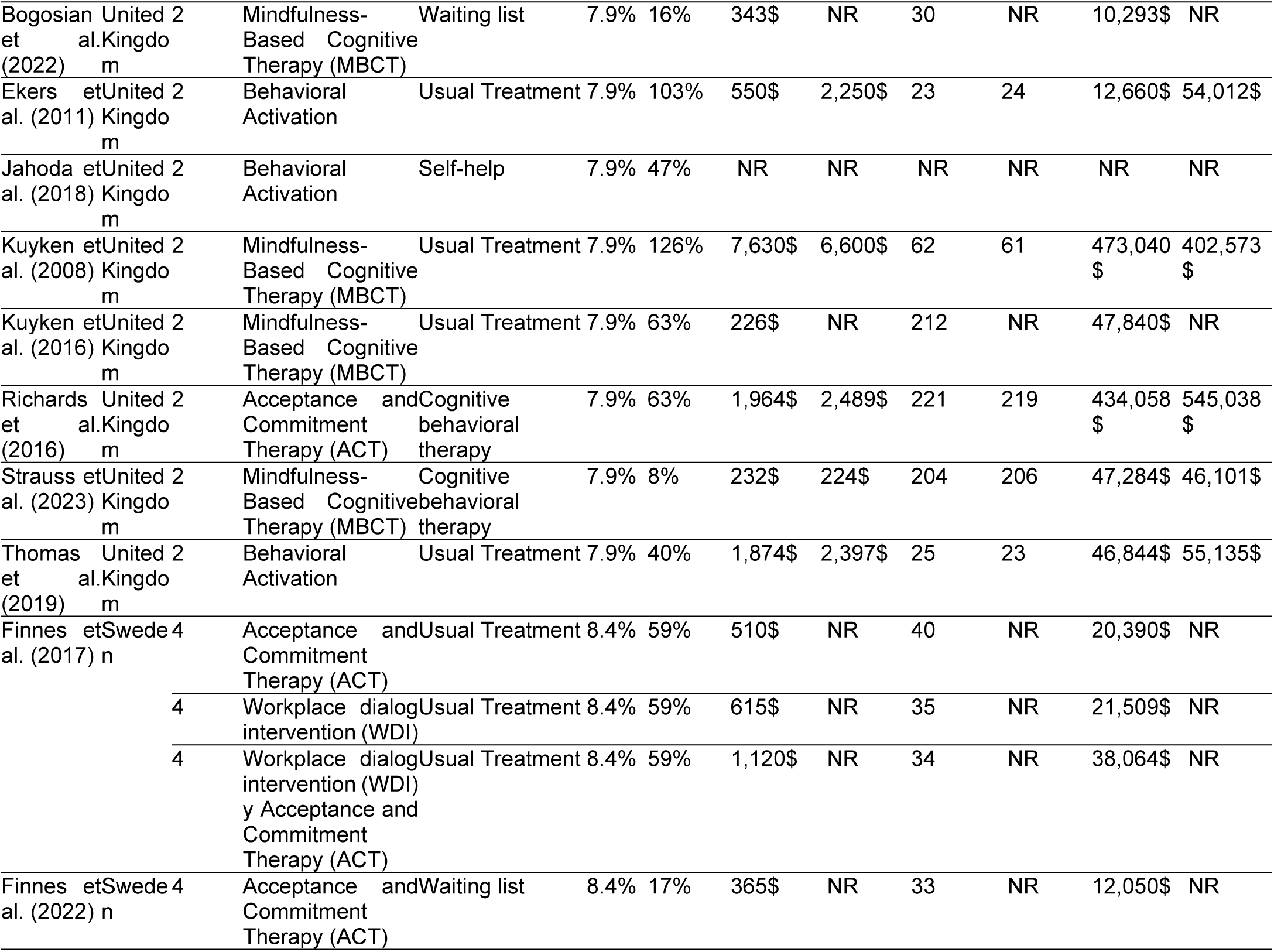
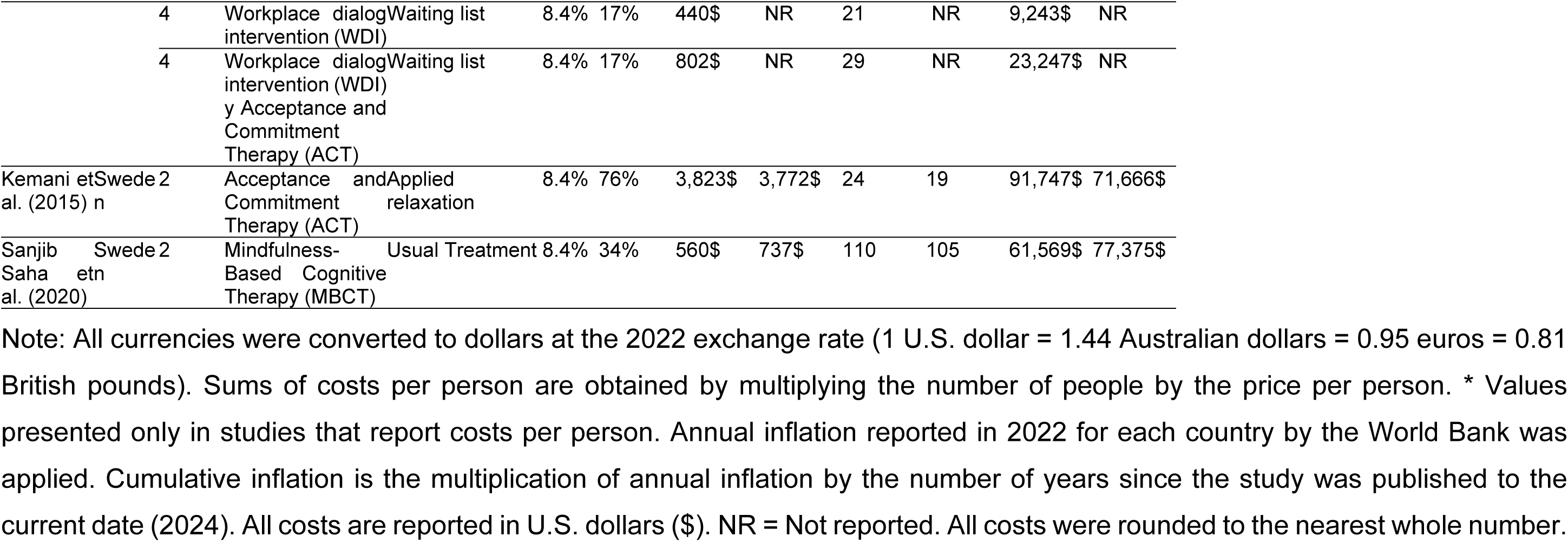
Calculation of costs per person, in the studies that report it for each arm. (n=26).

### Cost-effectiveness analysis

For the cost-effectiveness analysis, 10 out of 20 studies were meta-analyzed [27,28,31,32,36,38,40,42,43,45], including two studies with multiple intervention arms [27,36]. The other 10 studies were excluded due to insufficient data for meta-analysis. The general meta-analysis (Figure 3) showed that interventions based on third-generation therapies to treat symptoms of depression and anxiety were not significantly more effective in improving quality-adjusted life years (QALYs) compared to various control conditions (treatment as usual, waiting list, cognitive-behavioral therapy, and other interventions; Hedges’ g = 0.015, 95% CI: -0.001 to 0.032; P=0.070). Additionally, moderate heterogeneity was observed in the general meta-analysis (I²=47.8%, P=0.032). Also, the funnel plot identified that there was publication bias in the findings of the overall meta-analysis (Coeff=1.090, 95% CI: 0.309 to 1.870, P = 0.011). The funnel plot can be found in supplementary material 5.

**Figure 3.**
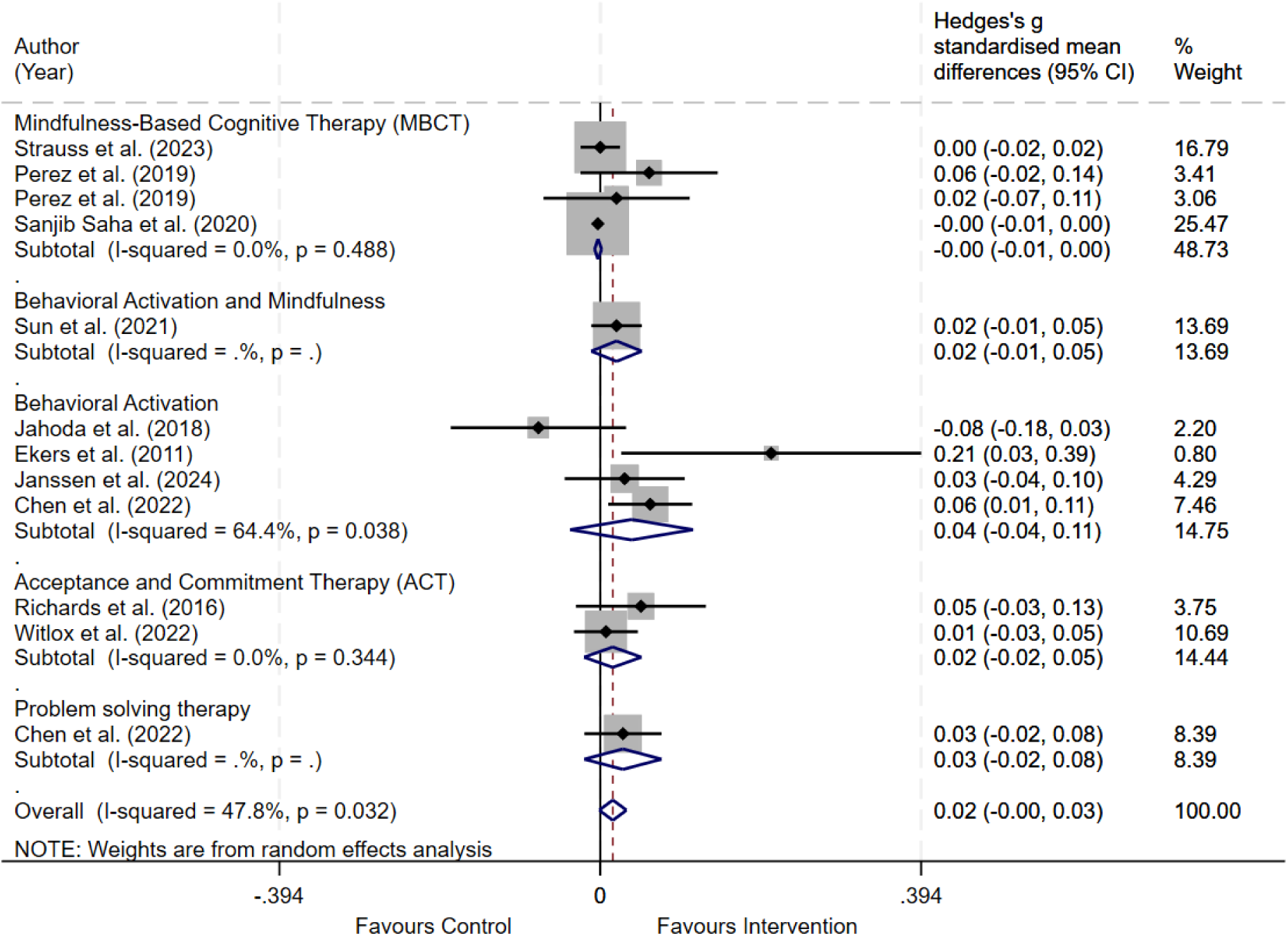
Meta-analysis by type of psychotherapy. Note: QALYs = quality-adjusted life years. The difference is calculated by subtracting the QALYs of the control group from the QALYs of the intervention group. All studies used the EQ-5D instrument in its different versions (EQ-5D-3L, EQ-5D-5L and EQ-5D-Y-3L) to measure QALYs.

A subgroup analysis was also performed according to the type of intervention, evaluating the results for QALYs. Mindfulness-Based Cognitive Therapy (MBCT), with three studies [31,36,38], did not show a significant effect compared to control conditions (g = -0.003, 95% CI: -0.007 to 0.002; P=0.221). Behavioral Activation and Mindfulness, represented by one study [43], also did not show a significant effect (g = 0.020, 95% CI: -0.011 to 0.051; P=0.207).

Behavioral Activation, with four studies [27,28,31,32], presented the highest heterogeneity (I² = 64.4%, P = 0.038) but did not show a significant effect (g = 0.039, 95% CI: -0.037 to 0.114; P=0.316). Acceptance and Commitment Therapy (ACT), with two studies [45,46], did not show a significant effect (g = 0.015, 95% CI: -0.020 to 0.051; P=0.393). Finally, Problem-Solving Therapy, represented by one study [27], also did not show a significant effect (g = 0.028, 95% CI: -0.020 to 0.075; P=0.249). It should be noted that it is not possible to assess heterogeneity in meta-analyses with only one study.

## Discussion

### Main findings

This systematic review and meta-analysis provides an updated and comprehensive assessment of the cost and cost-effectiveness of third-generation therapies for depressive and anxiety symptoms. Our findings significantly expand the existing knowledge in this field compared to the previous review [21], which analyzed 11 studies. Our study included 20 studies for cost analysis and 10 studies for cost-effectiveness analysis. Furthermore, while the previous study was limited to a descriptive and quality assessment using ROB [43], our study provides an evaluation of the risks of bias in economic analysis through ECOBIAS [23] and a cost-effectiveness meta-analysis based on random-effects models to control for the effect of heterogeneity across studies.

The cost analysis revealed that there was no significant difference in the average cost per person between groups receiving an intervention based on third-generation therapies (M = $2,494.6; SD = $7,090.9) and those assigned to a control condition (M = $1,912.0; SD = $2,081.3) (t = 0.253; p=0.802). This finding suggests that interventions based on third-generation therapies do not result in significant additional costs compared to control conditions, which is an important consideration for healthcare decision-makers.

Regarding the meta-analysis, which included 10 studies, it did not show a significant difference in QALY improvement between third-generation therapies and control conditions (Hedges’ g = 0.015, 95% CI: -0.001 to 0.032; P=0.070). This result contrasts with previous studies that have suggested greater effectiveness of these therapies [47]. The discrepancy could be attributed to several factors, including variability in therapy implementation, differences in study populations, or the limitations of QALYs in capturing the full benefits of mental health interventions [48].

It is important to note that specific meta-analyses were conducted for each therapy included in the study. Additionally, a global meta-analysis was performed, integrating all evaluated third-generation therapies. However, it must be acknowledged that these therapies are theoretically distinct and operate through different mechanisms of action, which should be considered when interpreting the findings. The global meta-analysis may exhibit high heterogeneity due to these conceptual differences, but it serves as a preliminary approach justified by the limited number of available studies.

Subgroup analyses based on specific third-generation therapies (MBCT, Behavioral Activation and Mindfulness, Behavioral Activation, ACT, and Problem-Solving Therapy) also did not yield significant effects on QALYs. However, these results should be interpreted cautiously due to the limited number of studies in each subgroup and the observed heterogeneity, particularly in the Behavioral Activation subgroup (I² = 64.4%, P = 0.038).

The risk of bias assessment using ECOBIAS showed that a significant proportion of the studies had a high risk of bias in several dimensions. Fifty percent of the studies showed a high risk of bias due to narrow perspective, which could lead to an underestimation of the total costs associated with the interventions. Forty-five percent presented a high risk of bias due to omission of cost measurement, which could affect the accuracy of cost-effectiveness estimates. Additionally, 46% did not report sufficient information on inappropriate discounting, which could influence the interpretation of long-term results [23].

These findings are consistent with other reviews in the mental health field, which have also found high levels of risk of bias [49,50]. This phenomenon could be explained by the inherent difficulties in psychotherapy research, such as the impossibility of complete blinding and variability in the implementation of interventions [51].

Although our findings suggest that third-generation therapies are not significantly more expensive than control conditions, the evidence on their cost-effectiveness is inconclusive. Health intelligence teams, especially in high-income countries, could consider implementing these therapies as part of usual treatment, but local studies are needed to confirm their cost-effectiveness in different contexts [48]. This consideration is particularly relevant given that 95% of the included studies were conducted in high-income countries, which limits the generalization of results to more resource-limited settings. Furthermore, the fact that 65% of the studies were conducted in populations with mental health problems underscores the importance of these therapies in treating psychological disorders, but also points to the need to investigate their efficacy in more diverse populations.

The lack of a significant difference in QALY improvement between third-generation therapies and control conditions raises questions about the justification for the additional costs associated with these interventions. However, it is important to consider that QALYs may not fully capture the benefits of these therapies, especially in the context of mental health. Health decision-makers must carefully weigh these findings along with other factors, such as the prevalence of depressive and anxiety symptoms in their population, resource availability, and patient preferences, when deciding on the implementation of these therapies [17].

### Strengths and limitations

A key strength of this study is its comprehensive nature, including a larger number of studies than previous reviews and employing both cost analysis and meta-analysis. The use of the ECOBIAS tool for assessing risk of bias in economic evaluations is another strength, providing a detailed understanding of the methodological quality of included studies.

However, our study has several limitations. An important limitation is the moderate heterogeneity observed in the general meta-analysis (I²=47.8%, P=0.032), which suggests variability among the included studies. This heterogeneity could be due to differences in the populations studied, implementation contexts, or variations in the application of therapies. Furthermore, the high heterogeneity observed in the Behavioral Activation subgroup indicates that the results for this particular therapy should be interpreted with caution. These variations limit our ability to make solid generalizations about the cost-effectiveness of third-generation therapies as a whole.

Another significant limitation is the high risk of bias observed in several included studies. Fifty percent of the studies presented a high risk of bias due to narrow perspective, and 45% due to omission of cost measurement. This could lead to an underestimation or overestimation of the actual costs associated with these therapies. Additionally, 46% of the studies did not report sufficient information on inappropriate discounting, which could affect the accuracy of long-term estimates.

Publication bias was also detected in our meta-analysis (Egger’s test: P = 0.011), which suggests that studies with positive results may be overrepresented in the literature. This could potentially lead to an overestimation of the effectiveness of these therapies.

To address these limitations, we propose improving the methodological quality of future studies by rigorously following established guidelines such as ECOBIAS [23]. This involves adopting a broader perspective in cost evaluation, ensuring a comprehensive measurement of all relevant costs, including indirect and social costs. It is also crucial to adequately report the discount rates used and justify their choice. Standardizing the implementation of third-generation therapies, to the extent possible, could help reduce heterogeneity between studies. Finally, the inclusion of robust sensitivity analyses would allow for the evaluation of the impact of different assumptions on cost-effectiveness results.

### Future Research Directions

Future research in this field should address several key areas to improve our understanding of the cost-effectiveness of third-generation therapies. First, it is crucial to expand research to more diverse contexts, particularly in low- and middle-income countries. This would allow for the evaluation of the cost-effectiveness of these therapies in different socioeconomic settings, addressing the current limitation that 95% of the included studies were conducted in high-income countries [48]. Additionally, it is important to explore outcomes other than QALYs that can more comprehensively capture the benefits of these therapies in mental health. This could include specific measures of depressive and anxiety symptoms, social and occupational functioning, and mental health-related quality of life.

Another interesting line of research would be the study of the cost-effectiveness of self-applied interventions based on third-generation therapies. These could offer a more accessible and scalable alternative, especially in resource-limited contexts. It would also be valuable to conduct long-term follow-up studies to evaluate the sustainability of the effects and costs associated with these therapies over time. Finally, future research should focus on the cost-effectiveness of these therapies in specific populations, such as older adults, adolescents, or people with medical comorbidities. This would allow for the identification of subgroups that may benefit more from these interventions and better guide the allocation of resources in mental health.

### Conclusions

This systematic review and meta-analysis provides a comprehensive assessment of the cost and cost-effectiveness of third-generation therapies for depressive and anxiety symptoms. Our findings indicate that these therapies do not entail significantly higher costs compared to control conditions. However, the meta-analysis did not show a significant difference in QALY improvement between third-generation therapies and control conditions. These results suggest that, despite their growing popularity, the cost-effectiveness of these therapies for the treatment of depressive and anxiety symptoms is not yet clearly established.

It is important to interpret these results in the context of the identified limitations, including heterogeneity between studies and the high risk of bias in several of them. Additionally, most studies were conducted in high-income countries, which limits the generalization of results to other contexts. More research is needed, especially in diverse settings and with more rigorous methodologies, to more precisely determine the value of these therapies in different mental health care contexts. Decision-makers should carefully consider these findings, along with other contextual factors and the specific needs of their population, when evaluating the implementation of these therapies in their health systems. As we move forward, it is crucial to continue investigating and refining our understanding of the cost-effectiveness of third-generation therapies to ensure that mental health resources are used in the most efficient and beneficial way possible.

## Acknowledgments

None.

## Data availability

Readers can access the full data of the study at: https://doi.org/10.6084/m9.figshare.26910076.v1

## Ethical approval and consent to participate

Not applicable.

## Consent to publication

Not applicable.

## Declaration of generative AI and AI-assisted technologies in the writing process

We used DeepL to translate specific sections of the manuscript and Grammarly to improve the wording of certain sections. All authors reviewed and approved the final version of the manuscript.

## Author Contributions

Yscenia Paredes-Gonzales: Conceptualization, Methodology, Validation, Formal analysis, Investigation, Data Curation, Writing - Original Draft, Visualization.

Gianfranco Centeno-Terrazas: Validation, Investigation, Writing - Original Draft, Writing - Review & Editing.

Kelly De la Cruz-Torralva: Validation, Data Curation. Elisa Romani-Huacani: Validation, Data Curation.

Yan Pieer: Validation, Data Curation.

David Villarreal-Zegarra: Conceptualization, Methodology, Validation, Formal analysis, Investigation, Data Curation, Writing - Original Draft, Visualization, Review & Editing, Supervision

## Supplementary Files

**Supplementary material 1.** PRISMA checklist.

Material suplementario 1. PRISMA checklist.
| Section and Topic | Item # | Checklist item | Location where item is reported |
| --- | --- | --- | --- |
| **TITLE** |  |  |  |
| Title | 1 | Identify the report as a systematic review. | pp. 1 |
| **ABSTRACT** |  |  |  |
| Abstract | 2 | See the PRISMA 2020 for Abstracts checklist. | pp. 5-6 |
| **INTRODUCTION** |  |  |  |
| Rationale | 3 | Describe the rationale for the review in the context of existing knowledge. | pp. 7-8 |
| Objectives | 4 | Provide an explicit statement of the objective(s) or question(s) the review addresses. | pp. 8 |
| **METHODS** |  |  |  |
| Eligibility criteria | 5 | Specify the inclusion and exclusion criteria for the review and how studies were grouped for the syntheses. | pp. 9 |
| Information sources | 6 | Specify all databases, registers, websites, organisations, reference lists and other sources searched or consulted to identify studies. Specify the date when each source was last searched or consulted. | pp. 10 |
| Search strategy | 7 | Present the full search strategies for all databases, registers and websites, including any filters and limits used. | pp. 47 - 51, Supplementary material 2 |
| Selection process | 8 | Specify the methods used to decide whether a study met the inclusion criteria of the review, including how many reviewers screened each record and each report retrieved, whether they worked independently, and if applicable, details of automation tools used in the process. | pp. 10 |
| Data collection process | 9 | Specify the methods used to collect data from reports, including how many reviewers collected data from each report, whether they worked independently, any processes for obtaining or confirming data from study investigators, and if applicable, details of automation tools used in the process. | pp. 10-11 |
| Data items | 10a | List and define all outcomes for which data were sought. Specify whether all results that were compatible with each outcome domain in each study were sought (e.g. for all measures, time points, analyses), and if not, the methods used to decide which results to collect. | pp. 10-11 |
|  | 10b | List and define all other variables for which data were sought (e.g. participant and intervention characteristics, funding sources). Describe any assumptions made about any missing or unclear information. | pp. 10 - 11 |
| Study risk of bias assessment | 11 | Specify the methods used to assess risk of bias in the included studies, including details of the tool(s) used, how many reviewers assessed each study and whether they worked independently, and if applicable, details of automation tools used in the process. | pp. 11 |
| Effect measures | 12 | Specify for each outcome the effect measure(s) (e.g. risk ratio, mean difference) used in the synthesis or presentation of results. | pp. 12 |
| Synthesis methods | 13a | Describe the processes used to decide which studies were eligible for each synthesis (e.g. tabulating the study intervention characteristics and comparing against the planned groups for each synthesis (item #5)). | pp. 12-13 |
|  | 13b | Describe any methods required to prepare the data for presentation or synthesis, such as handling of missing summary statistics, or data conversions. | pp. 12-13 |
|  | 13c | Describe any methods used to tabulate or visually display results of individual studies and syntheses. | pp. 12-13 |
|  | 13d | Describe any methods used to synthesize results and provide a rationale for the choice(s). If meta-analysis was performed, describe the model(s), method(s) to identify the presence and extent of statistical heterogeneity, and software package(s) used. | pp. 12-13 |
|  | 13e | Describe any methods used to explore possible causes of heterogeneity among study results (e.g. subgroup analysis, meta-regression). | pp. 13 |
|  | 13f | Describe any sensitivity analyses conducted to assess robustness of the synthesized results. | pp. |
| Reporting bias assessment | 14 | Describe any methods used to assess risk of bias due to missing results in a synthesis (arising from reporting biases). | pp. 13 |
| Certainty assessment | 15 | Describe any methods used to assess certainty (or confidence) in the body of evidence for an outcome. | No applicable |
| **RESULTS** |  |  |  |
| Study selection | 16a | Describe the results of the search and selection process, from the number of records identified in the search to the number of studies included in the review, ideally using a flow diagram. | pp. 14-15 |
|  | 16b | Cite studies that might appear to meet the inclusion criteria, but which were excluded, and explain why they were excluded. | pp. 52-59, Supplementary material 3 |
| Study characteristics | 17 | Cite each included study and present its characteristics. | pp. 16-18 |
| Risk of bias in studies | 18 | Present assessments of risk of bias for each included study. | pp. 18-20 |
| Results of individual studies | 19 | For all outcomes, present, for each study: (a) summary statistics for each group (where appropriate) and (b) an effect estimate and its precision (e.g. confidence/credible interval), ideally using structured tables or plots. | pp. 59-62, Supplementary material 4 |
| Results of syntheses | 20a | For each synthesis, briefly summarise the characteristics and risk of bias among contributing studies. | pp. 21 |
|  | 20b | Present results of all statistical syntheses conducted. If meta-analysis was done, present for each the summary estimate and its precision (e.g. confidence/credible interval) and measures of statistical heterogeneity. If comparing groups, describe the direction of the effect. | pp. 30-31 |
|  | 20c | Present results of all investigations of possible causes of heterogeneity among study results. | pp. 30-31 |
|  | 20d | Present results of all sensitivity analyses conducted to assess the robustness of the synthesized results. | pp. 12 |
| Reporting biases | 21 | Present assessments of risk of bias due to missing results (arising from reporting biases) for each synthesis assessed. | pp. 63, Supplementary material 5 |
| Certainty of evidence | 22 | Present assessments of certainty (or confidence) in the body of evidence for each outcome assessed. | No applicable |
| DISCUSSION |  |  |  |
| Discussion | 23a | Provide a general interpretation of the results in the context of other evidence. | pp. 32-33 |
|  | 23b | Discuss any limitations of the evidence included in the review. | pp. 34 |
|  | 23c | Discuss any limitations of the review processes used. | pp. 34 |
|  | 23d | Discuss implications of the results for practice, policy, and future research. | pp. 35 |
| OTHER INFORMATION |  |  |  |
| Registration and protocol | 24a | Provide registration information for the review, including register name and registration number, or state that the review was not registered. | pp. 9 |
|  | 24b | Indicate where the review protocol can be accessed, or state that a protocol was not prepared. | pp. 9 |
|  | 24c | Describe and explain any amendments to information provided at registration or in the protocol. | No applicable |
| Support | 25 | Describe sources of financial or non-financial support for the review, and the role of the funders or sponsors in the review. | pp. 3 |
| Competing interests | 26 | Declare any competing interests of review authors. | pp. 3 |
| Availability of data, code and other materials | 27 | Report which of the following are publicly available and where they can be found: template data collection forms; data extracted from included studies; data used for all analyses; analytic code; any other materials used in the review. | pp. 3 |

**Supplementary table 2.**
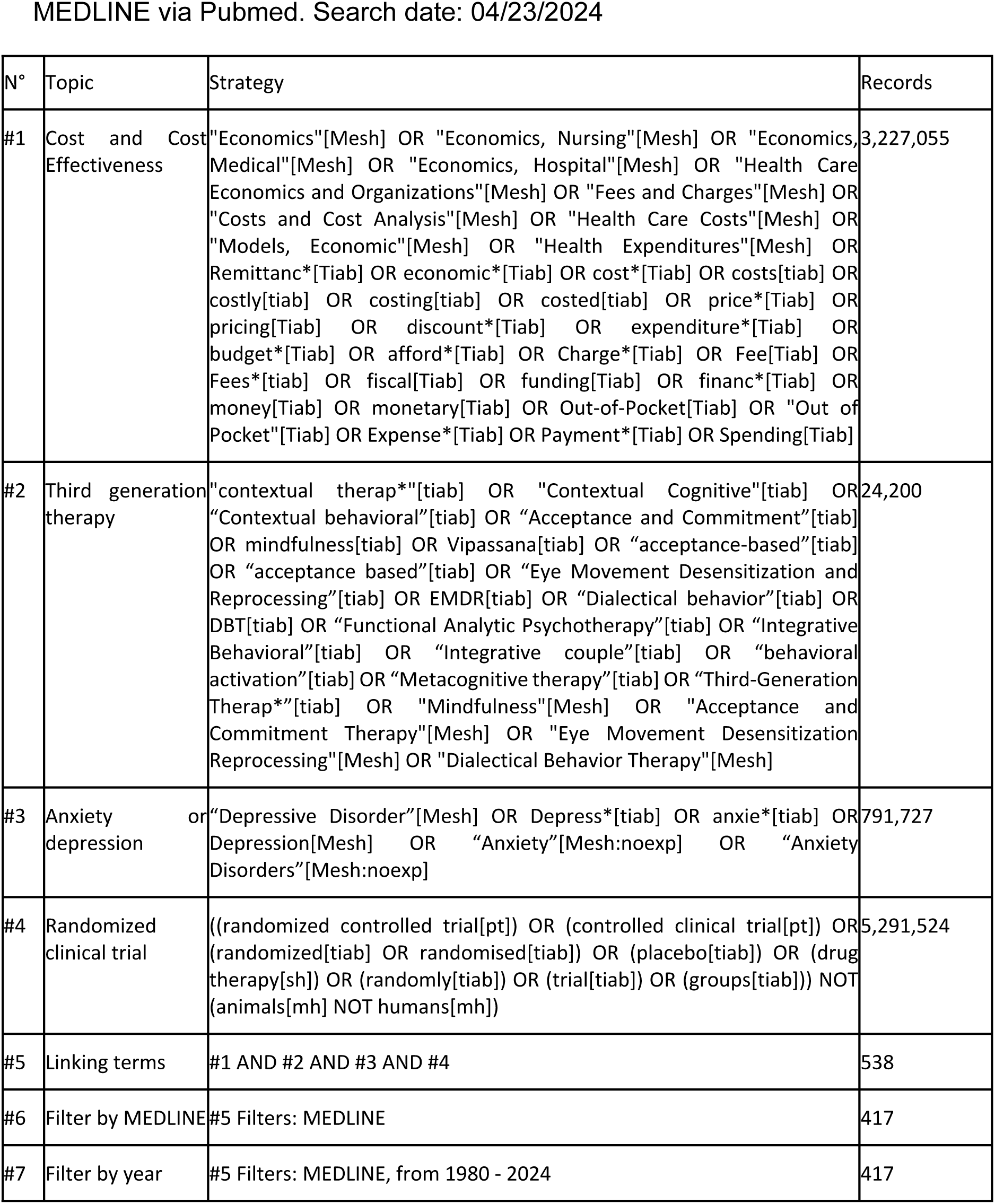
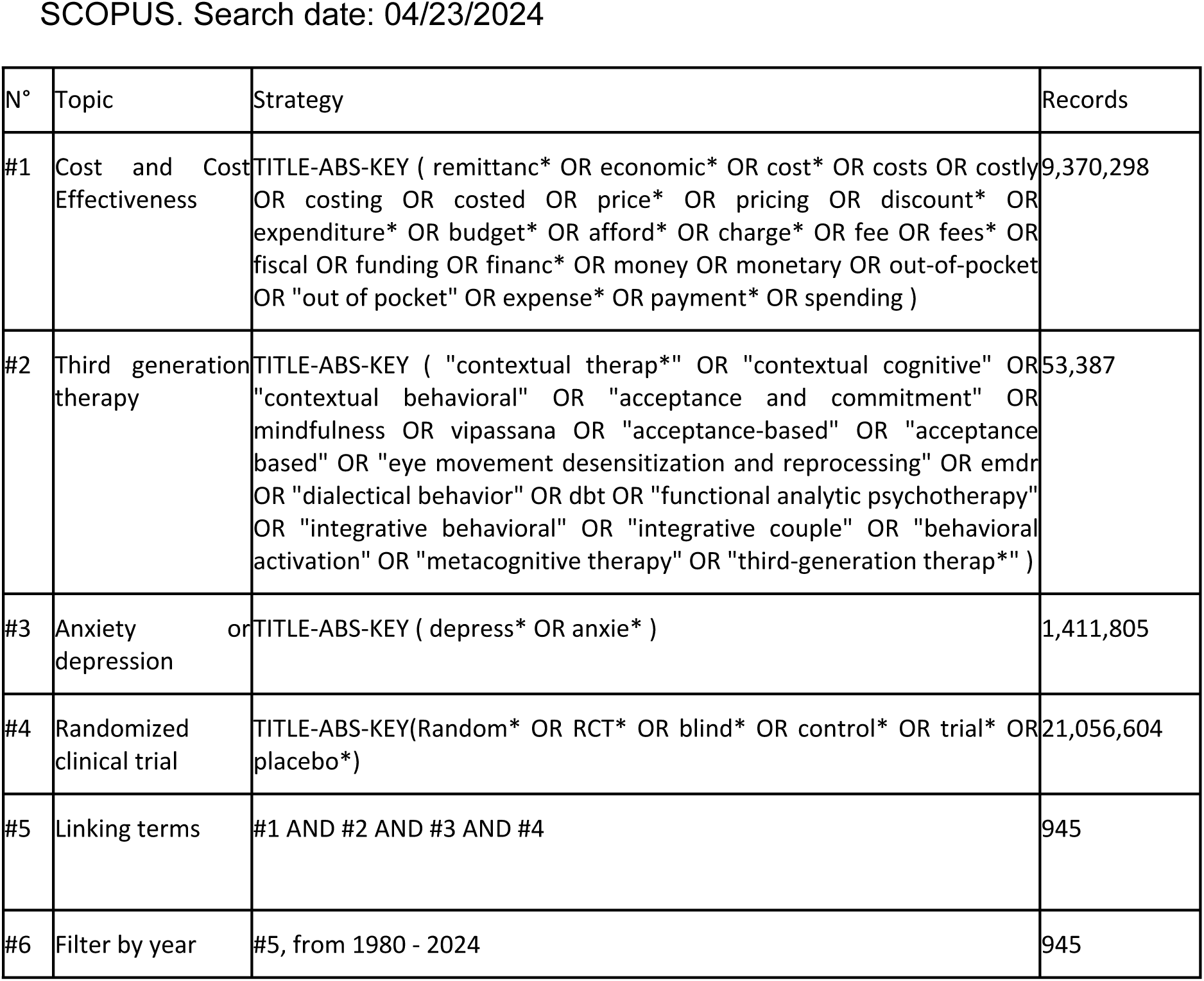
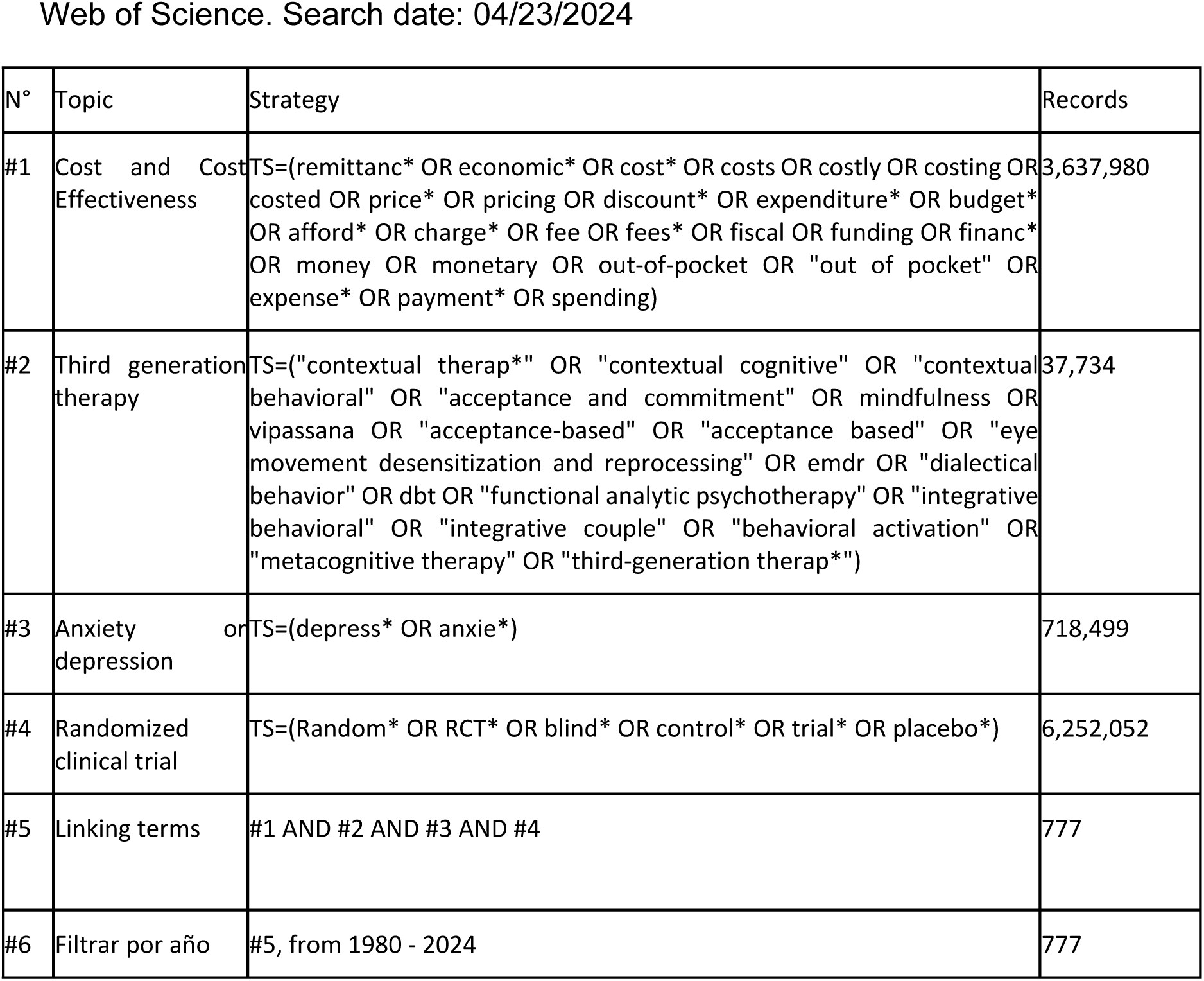
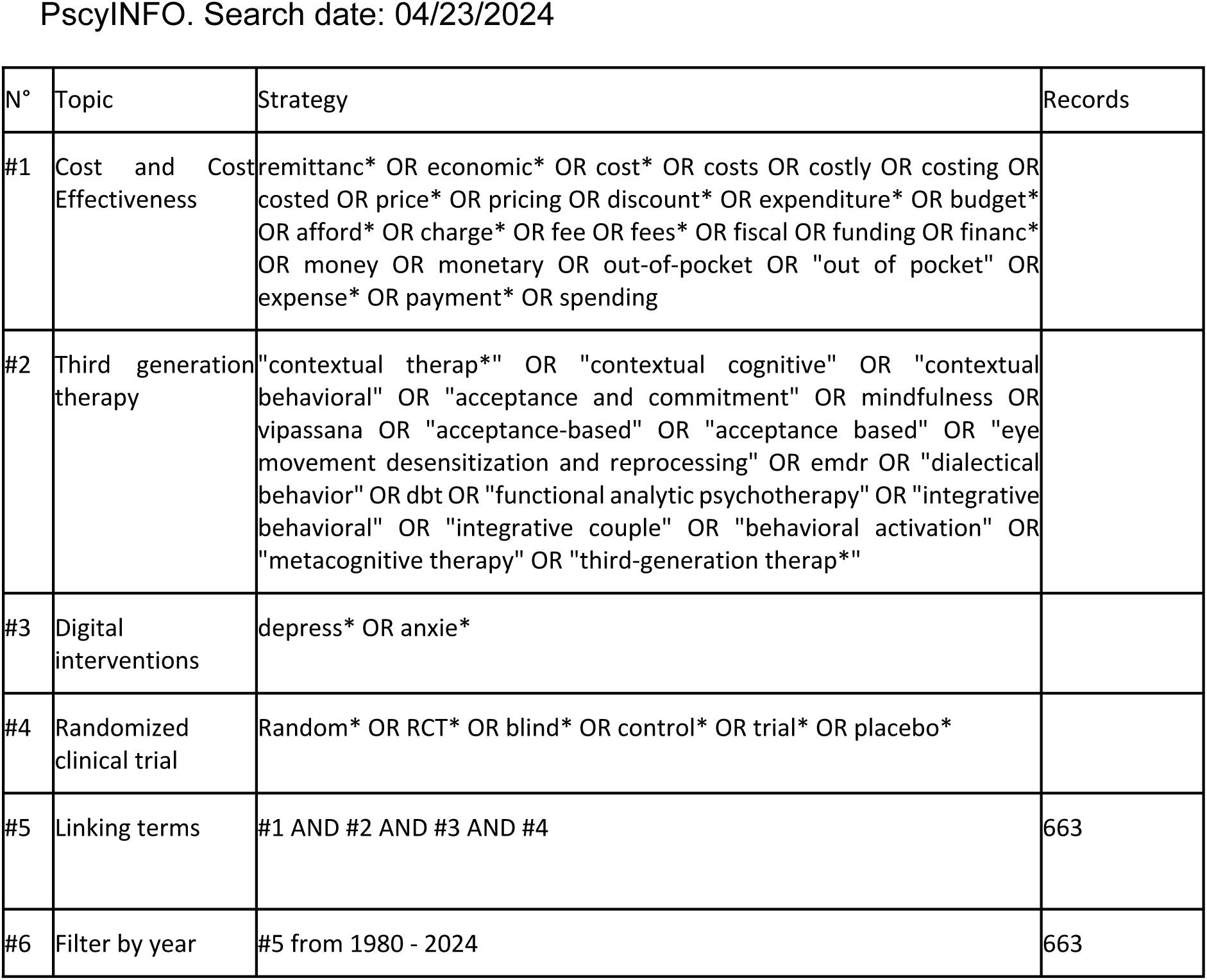
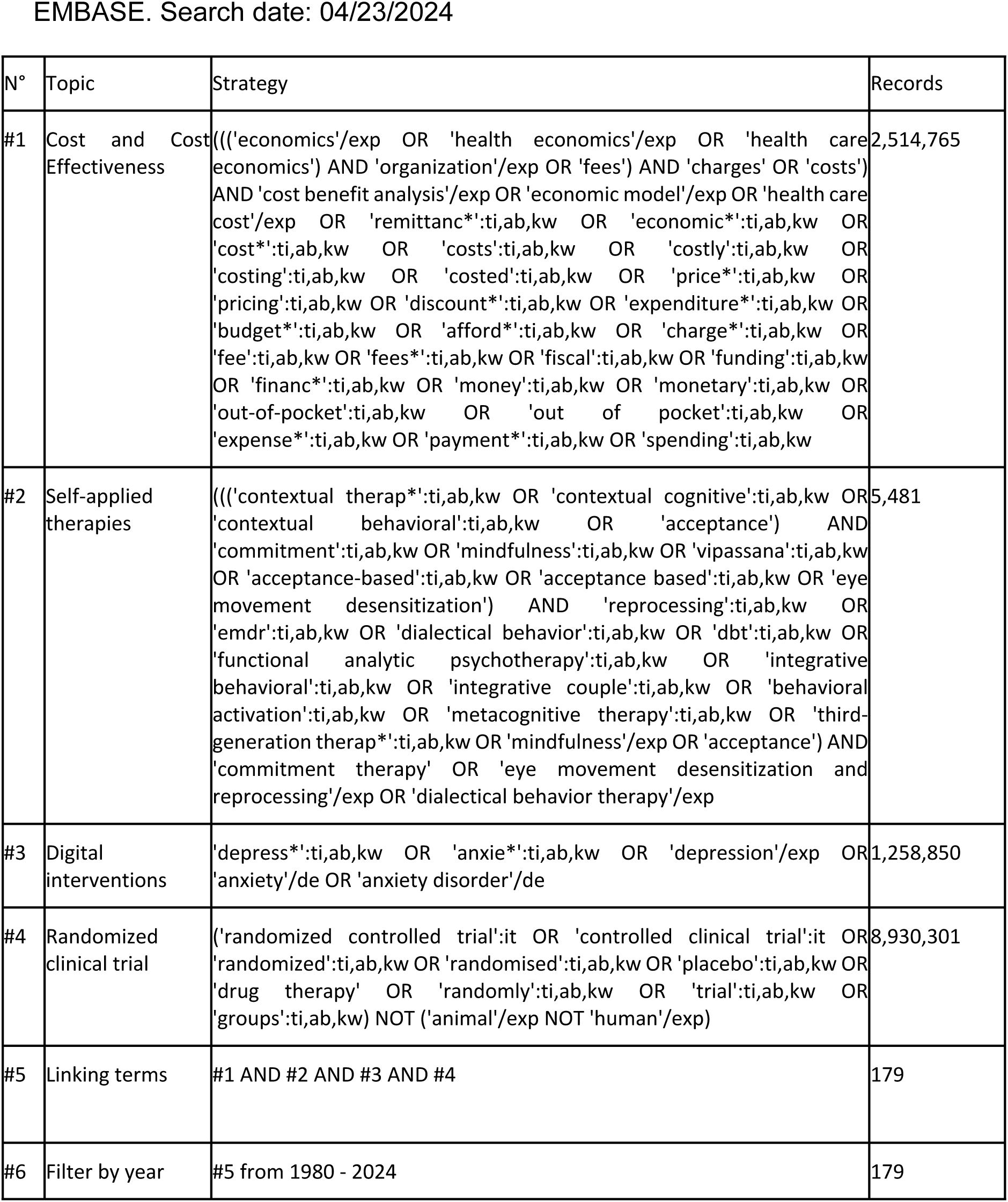
Search strategies.

**Supplementary table 3.**
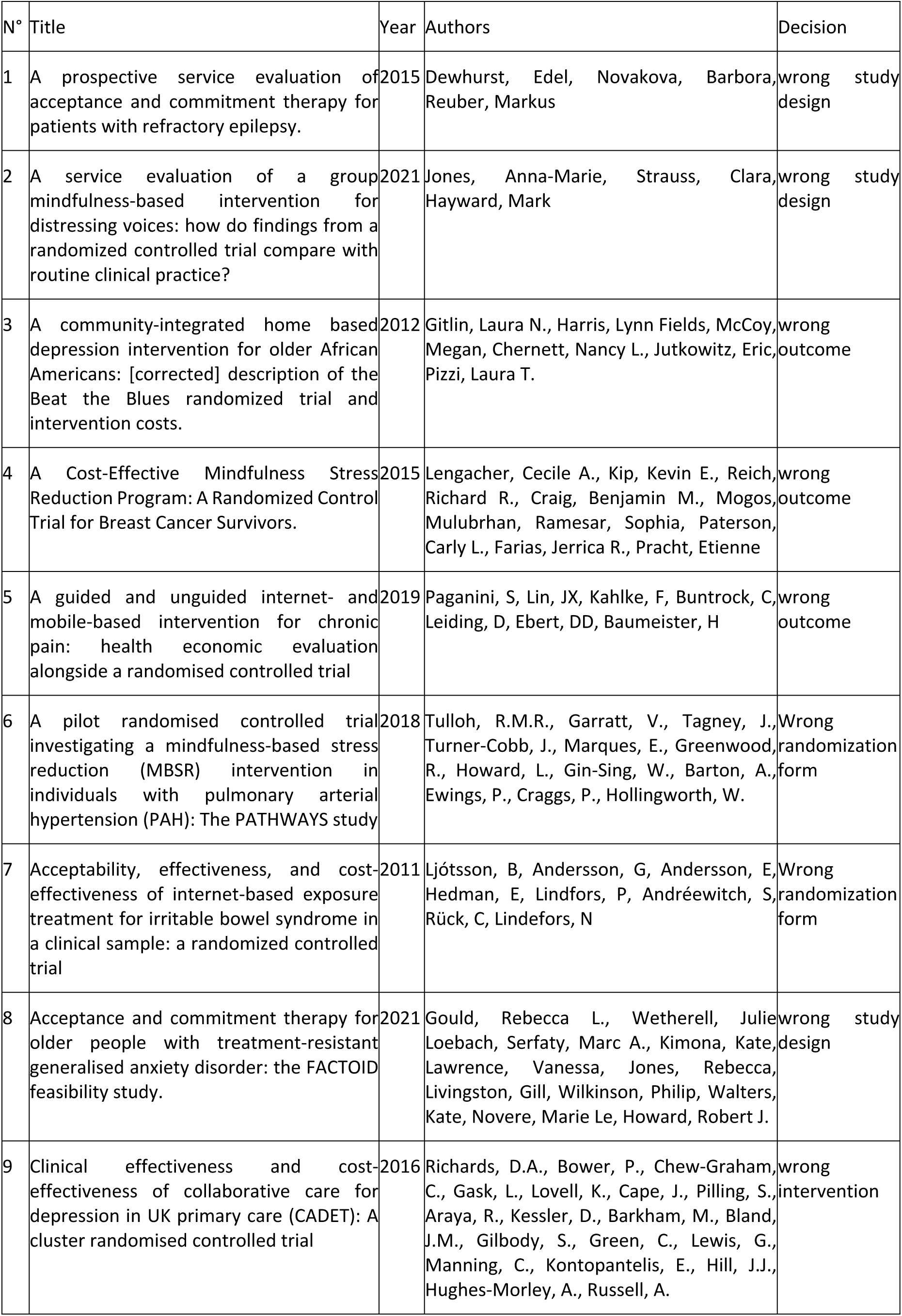
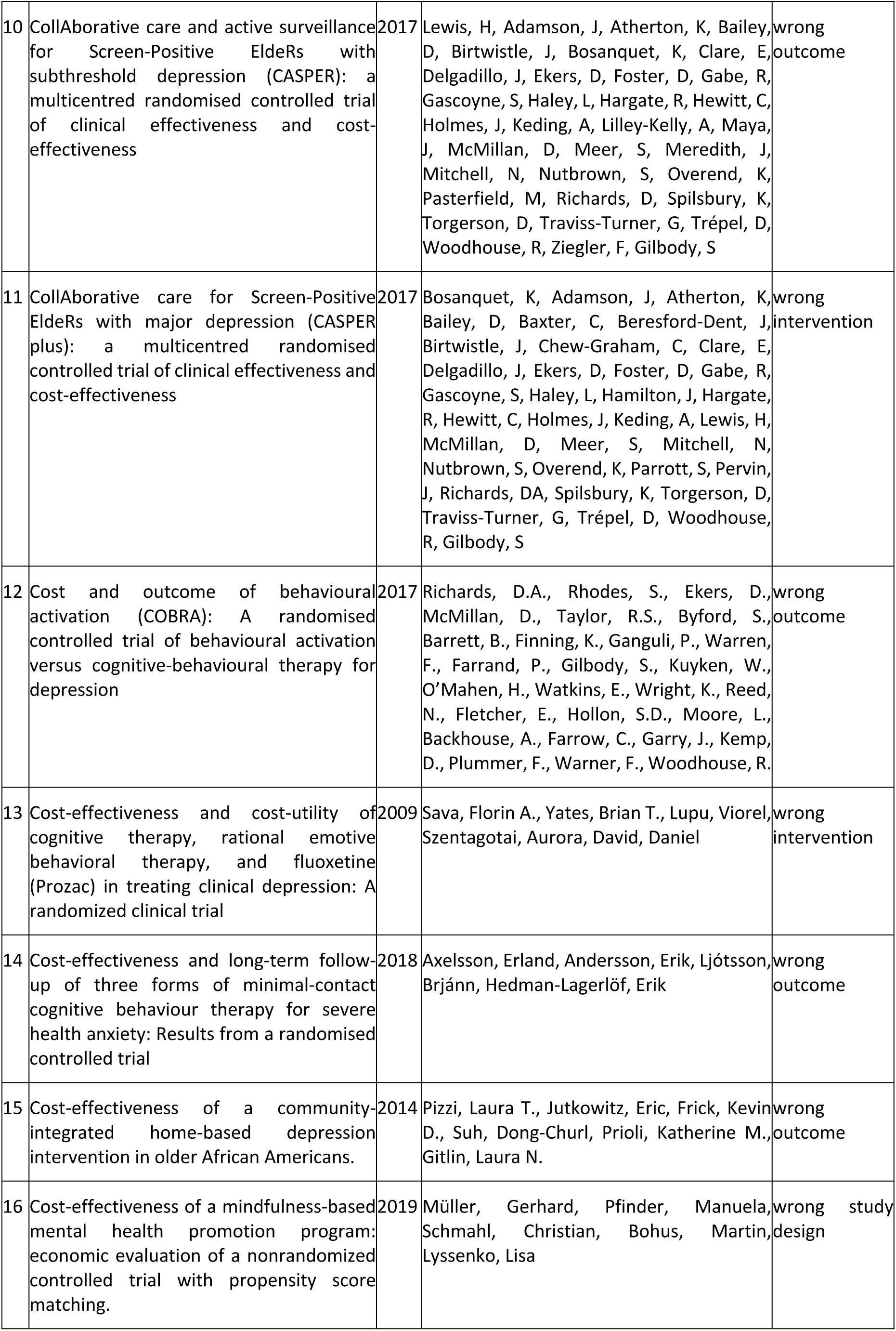
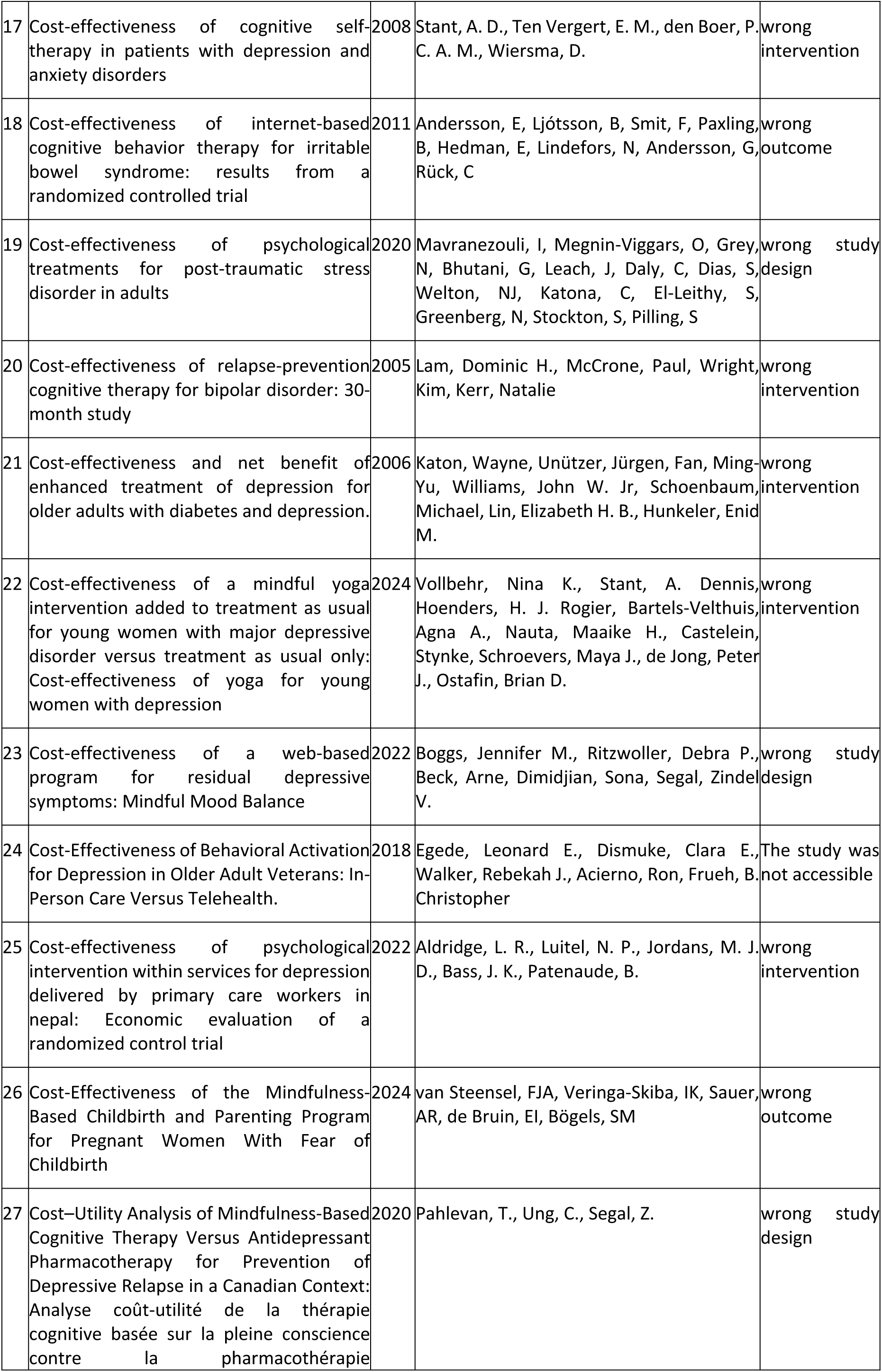
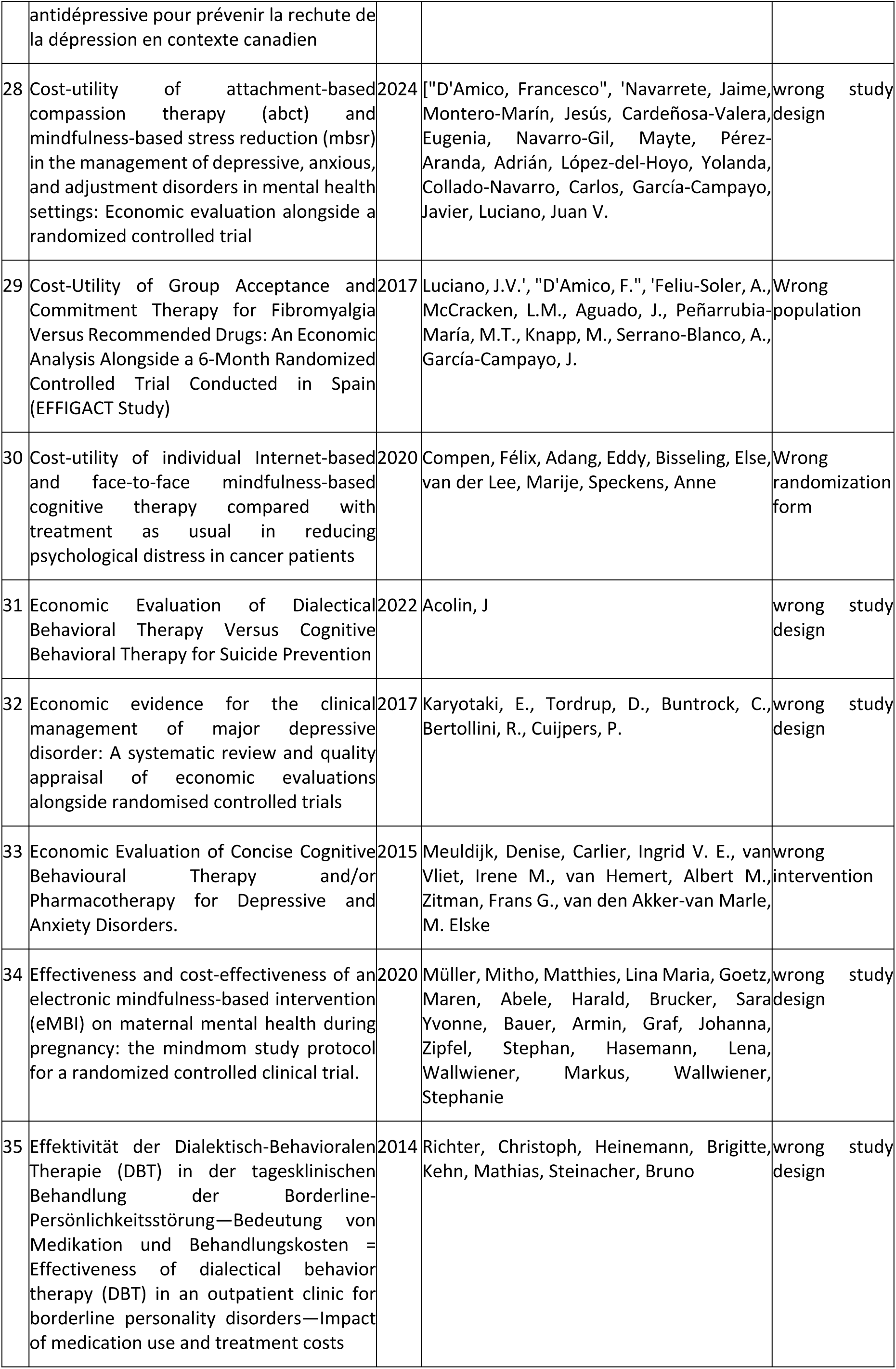
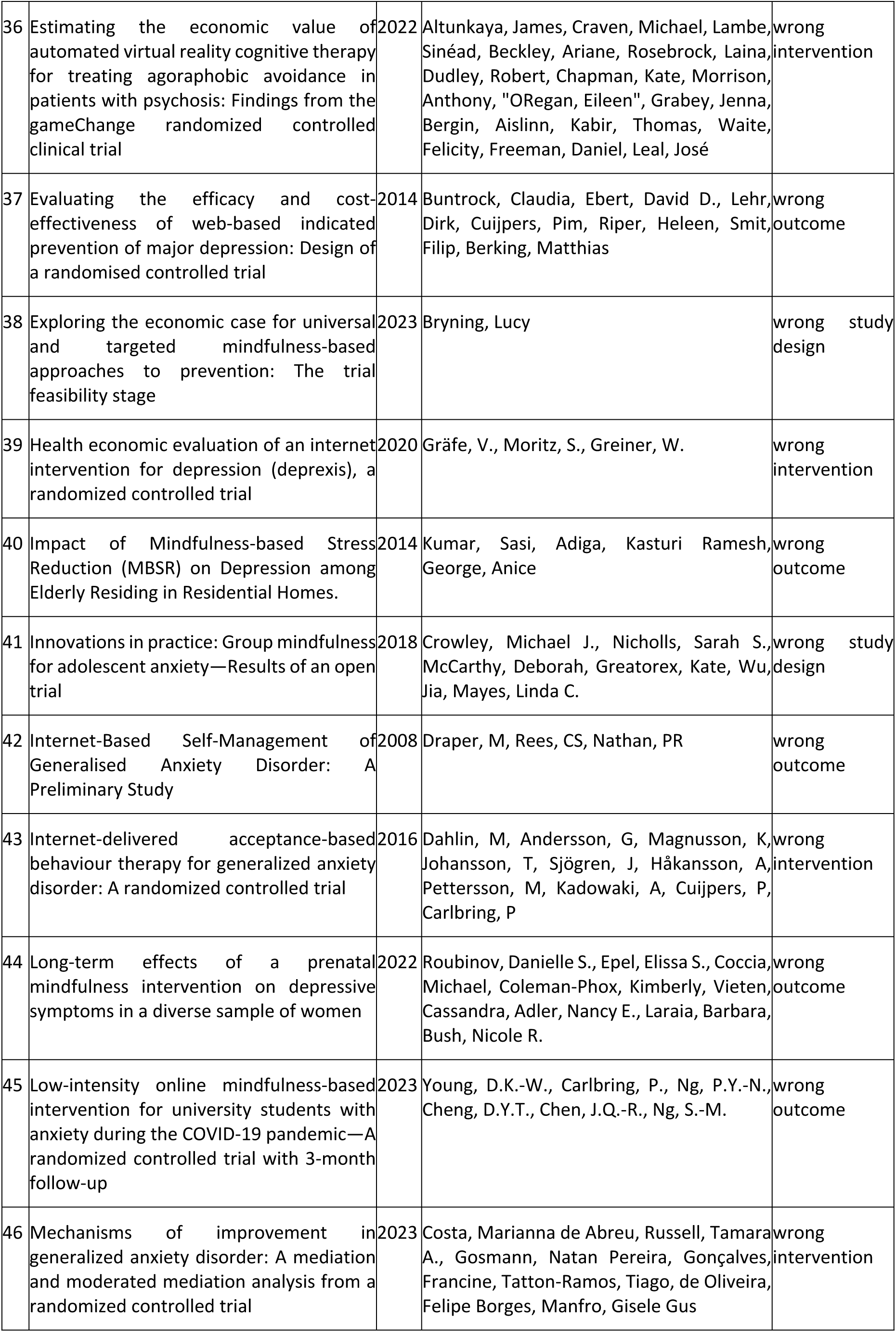
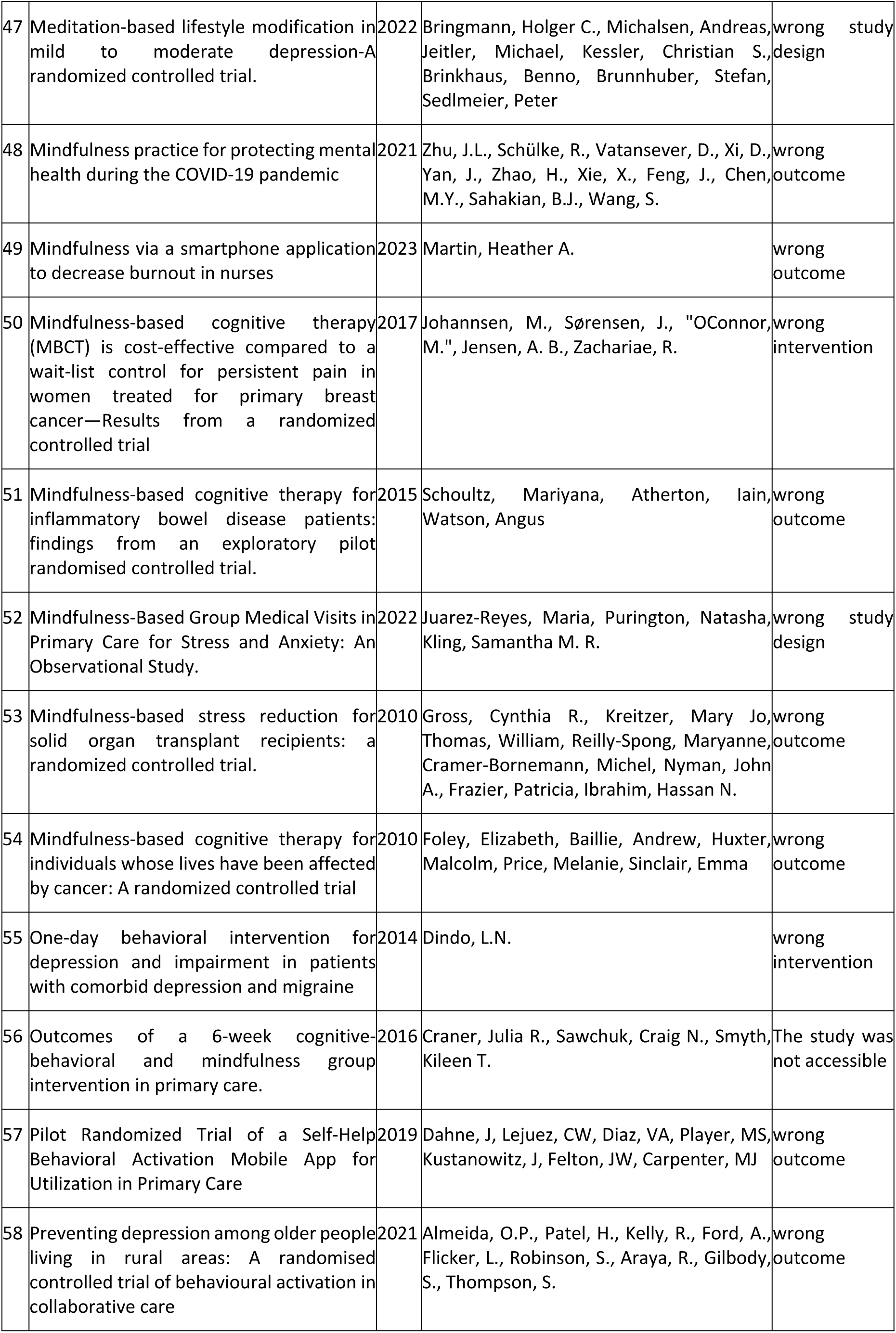
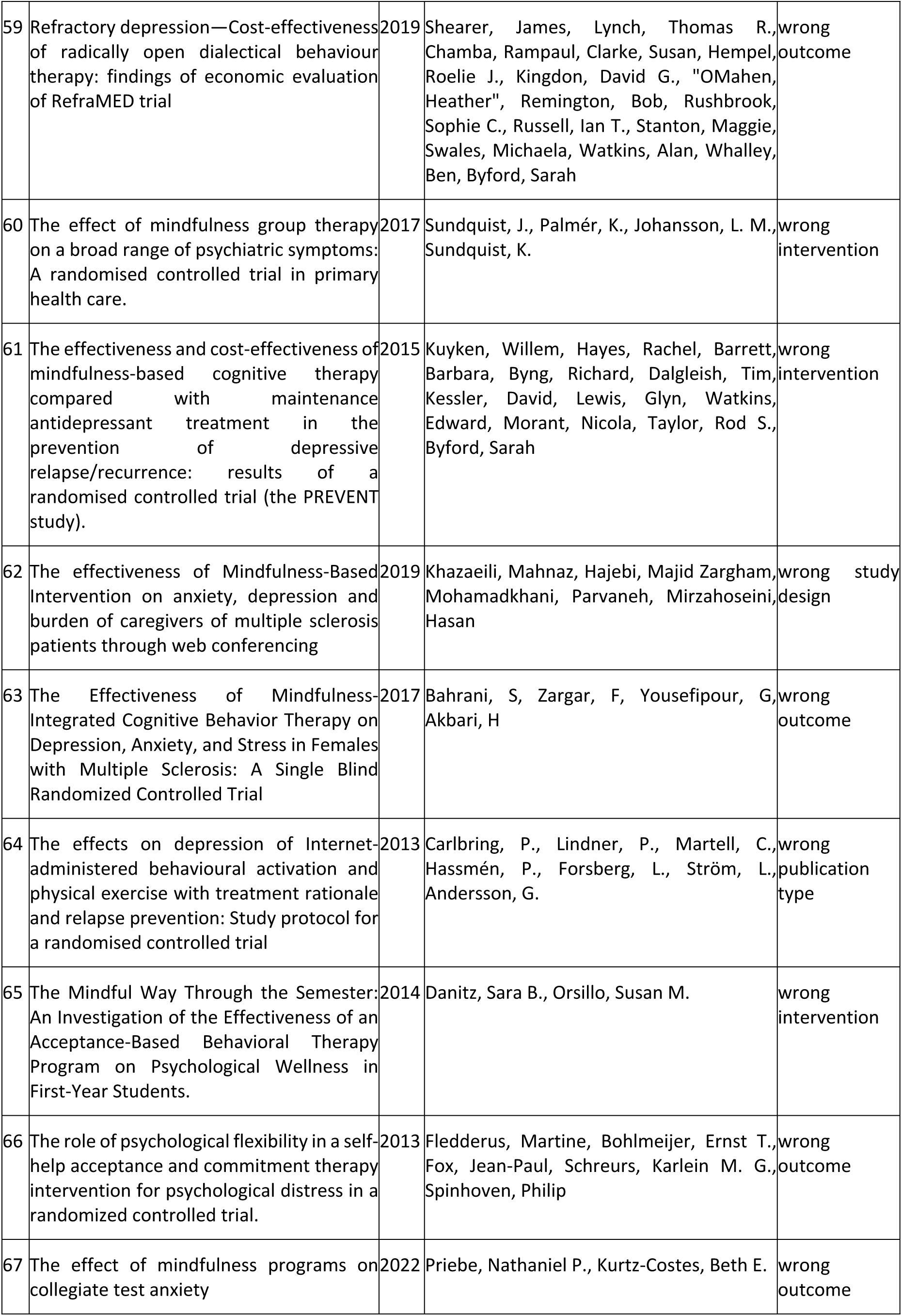
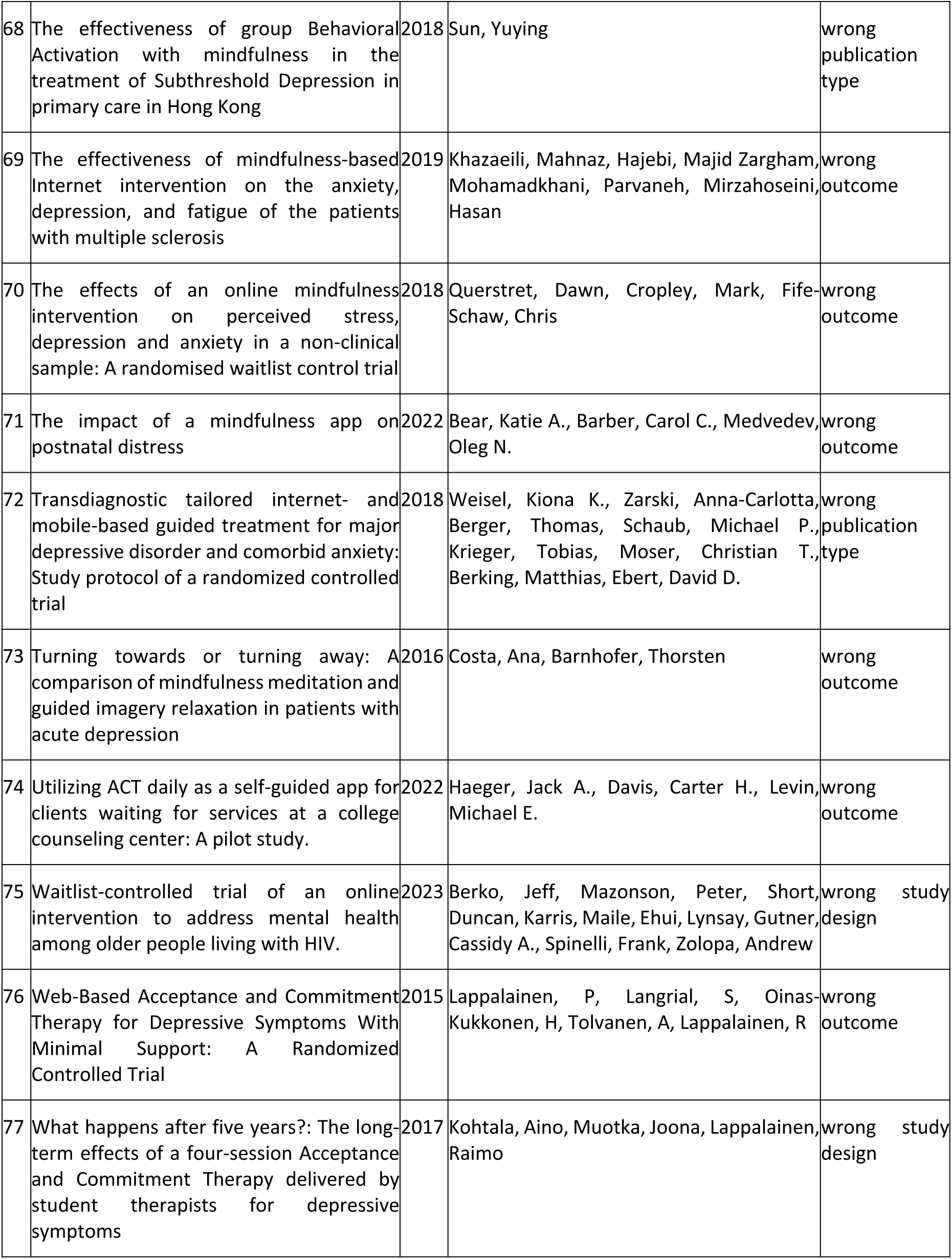
Records excluded by full text (n=77).

**Supplementary table 4.**
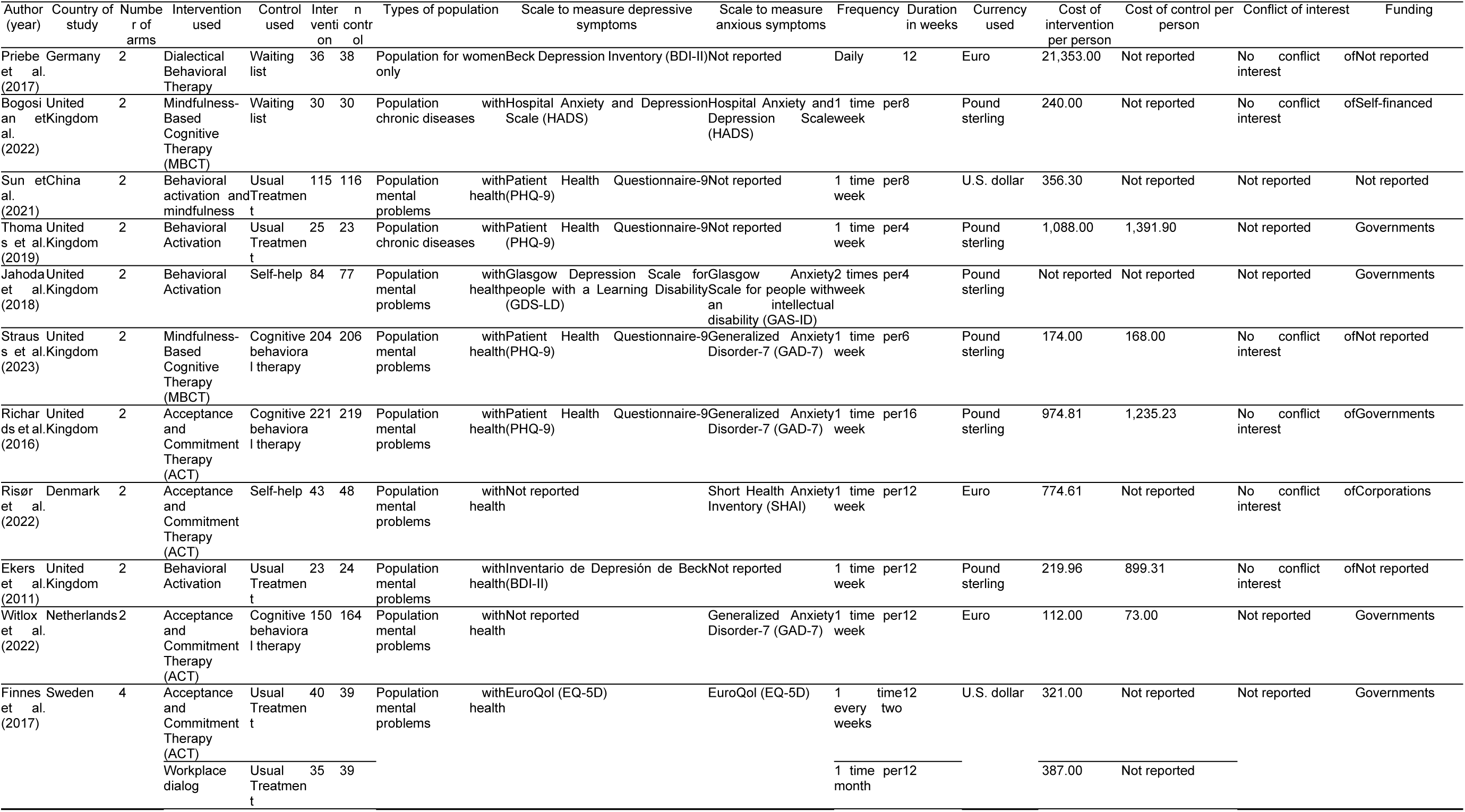
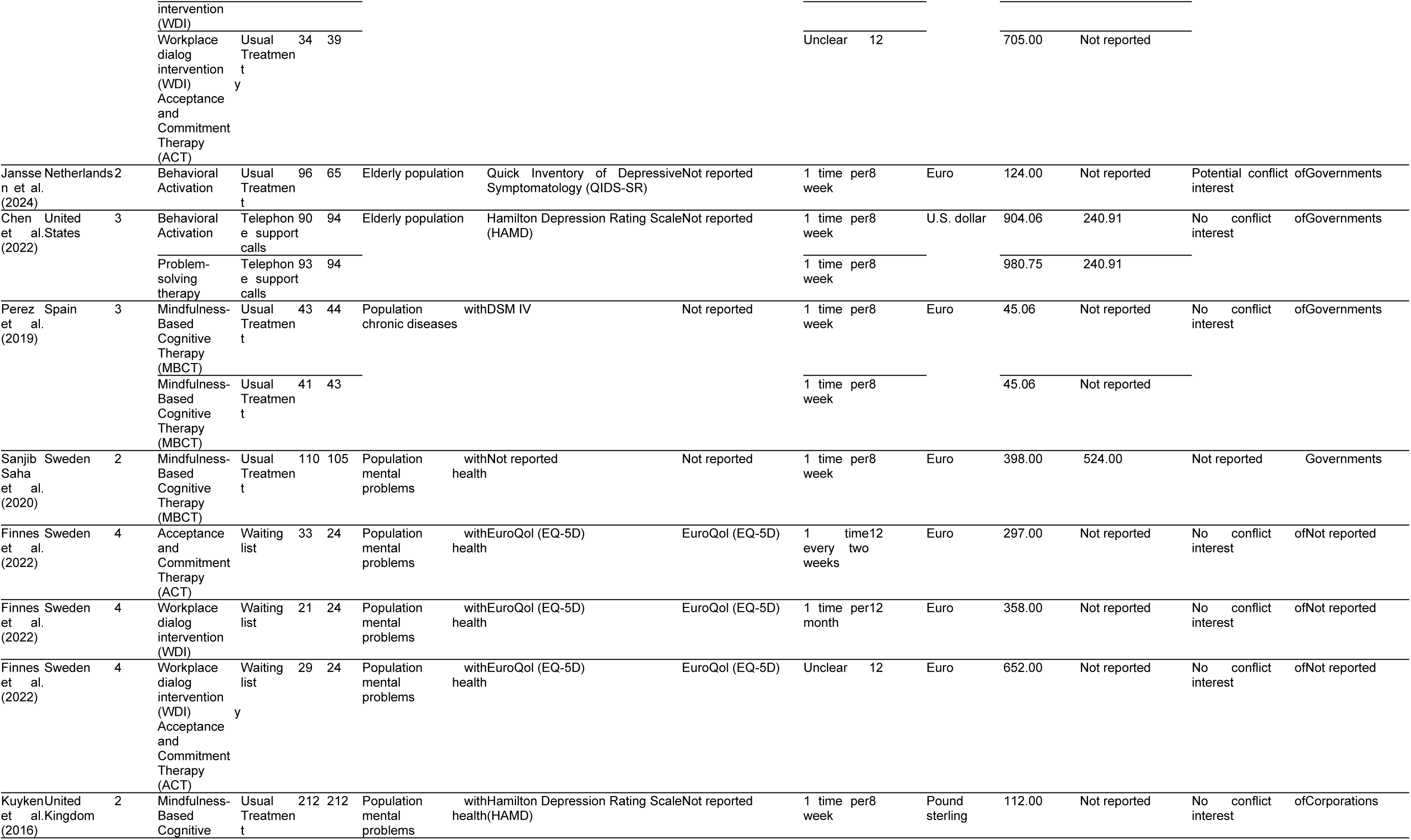
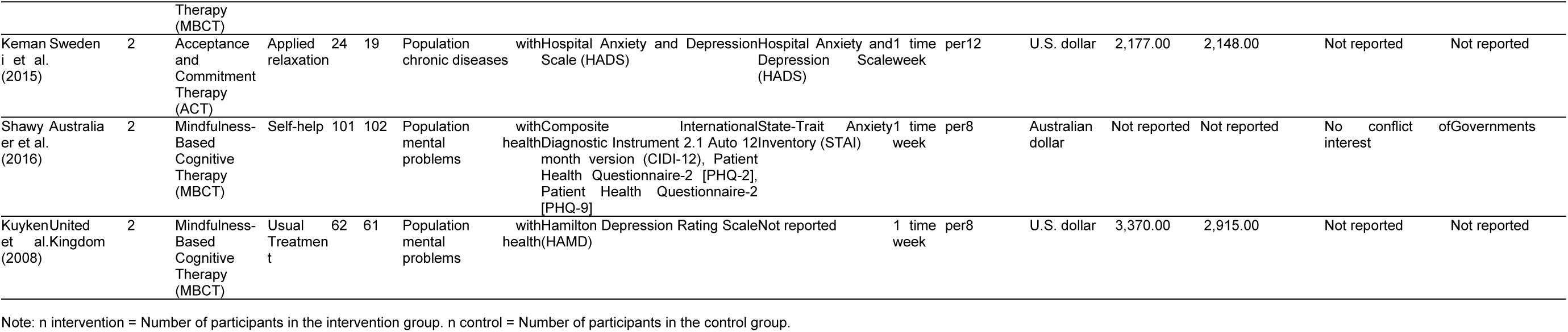
Individual description of studies (n=20).

**Supplementary table 5.**
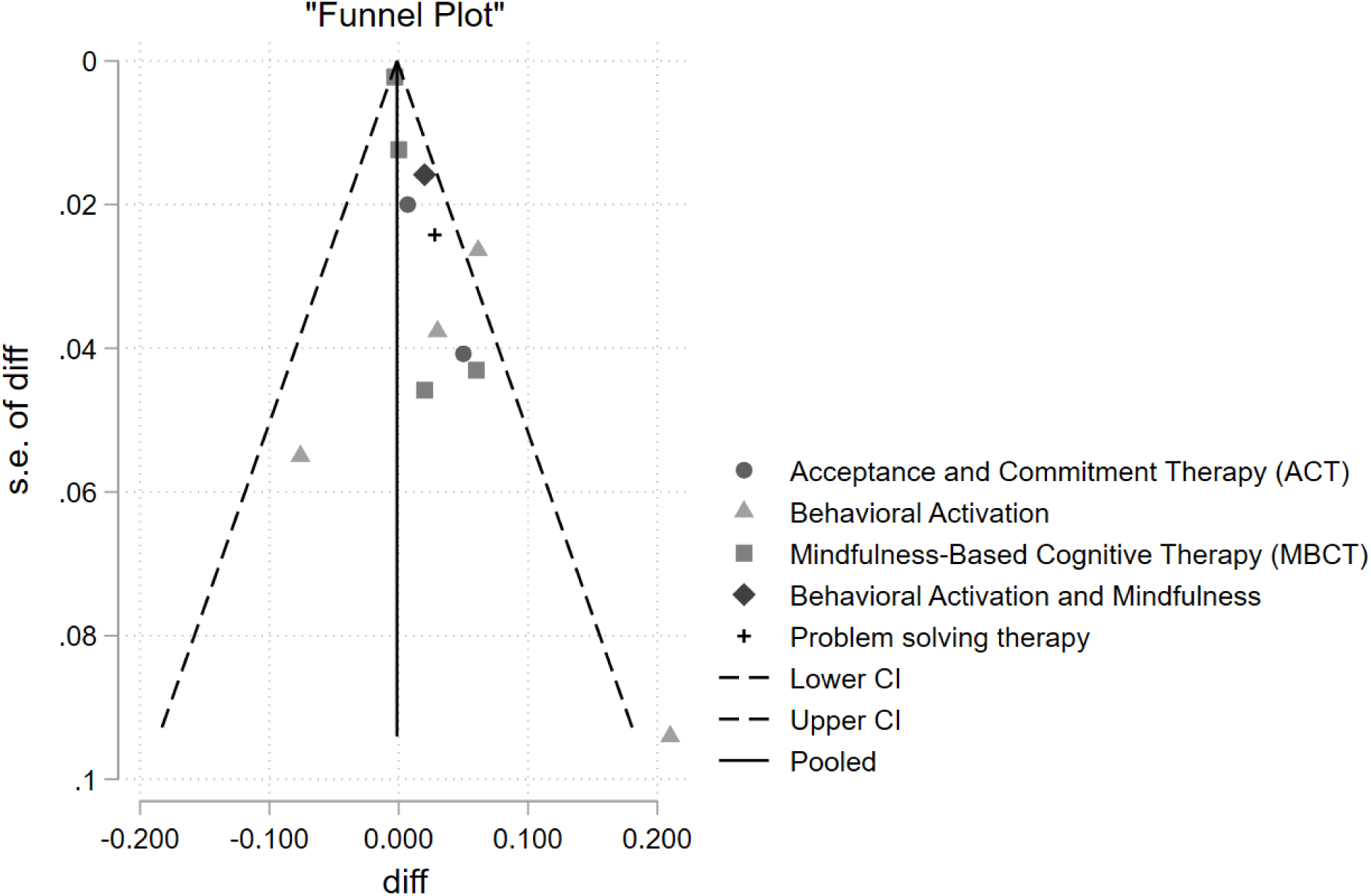
Publication bias analysis of the QALYS meta-analysis (n=10).

